# Semaglutide alters the human embryo-endometrium interface

**DOI:** 10.64898/2026.03.03.26347354

**Authors:** Apostol Apostolov, Amruta D. S. Pathare, Darja Lavogina, Cheng Zhao, Keiu Kask, Lucia Blanco-Rodriguez, Susana Ruiz-Durán, Sanjiv Risal, Ilmatar Rooda, Pauliina Damdimopoulou, Merli Saare, Maire Peters, Hannu Koistinen, Ganesh Acharya, Masoud Zamani Esteki, Fredrik Lanner, Alberto Sola-Leyva, Andres Salumets

**Author notes:** Correspondence to: Andres Salumets. These authors contributed equally (Apostol Apostolov, Amruta D. S. Pathare, Alberto Sola-Leyva). Co-last authors (Fredrik Lanner, Alberto Sola-Leyva, Andres Salumets).

## Abstract

The use of semaglutide (SE), a glucagon-like peptide-1 receptor agonist (GLP-1RA) with glucose-lowering and weight-loss effects, has risen rapidly, particularly among women of reproductive age. While preclinical studies suggest benefits for ovarian function via the hypothalamic–pituitary–ovarian axis, its impact on the endometrial–embryo interface remains unclear. Here, we show that GLP-1R is dynamically expressed in human endometrium, restricted to epithelial cells and markedly upregulated during the mid-secretory phase of the menstrual cycle. SE activated intracellular cAMP signalling and enhanced epithelial metabolism, driving a shift toward oxidative phosphorylation. Notably, even in the absence of steroid hormone priming, the highest SE concentration upregulates several endometrial receptivity markers, whereas under hormonal priming, the same concentration modestly reduces expression of a key receptivity marker, PAEP/glycodelin. By contrast, in stromal cells lacking detectable GLP-1R, SE disrupts decidualization, induces endoplasmic reticulum stress and suppresses cell-cycle at G2/M phase. Human embryo models, blastoids, expressed GLP-1R and underwent concordant SE-mediated transcriptional remodeling in epiblast and trophectoderm lineages, encompassing changes in metabolism and epigenetic regulation, but without shifts in lineage proportions. Notably, SE restored blastoid attachment under hormone-deprived conditions at all tested concentrations, but did not further enhance attachment when physiological hormonal priming was already present. Together, these findings reveal a compartment-specific mismatch, as SE augments epithelial and embryonic metabolic activity but compromises stromal support for implantation, with potential consequences for implantation due to stromal dysfunction.

## Introduction

Initially developed as glucose-lowering drugs, glucagon-like peptide-1 receptor agonists (GLP-1RAs) have emerged as multipurpose therapeutics, extending their reach from type 2 diabetes to obesity and liver disease^1,2^. Their clinical efficacy is rooted in activation of the GLP-1R, a class B G protein–coupled receptor that signals primarily through the canonical Gs–adenylyl cyclase–cAMP pathway, engaging protein kinase A (PKA) and EPAC2 to enhance calcium influx and stimulate glucose-dependent insulin secretion^3,4^. Obesity is one of the major contributors to female infertility, affecting approximately 20–25% of reproductive-age women globally^5^, exerting detrimental effects on reproductive health through endocrine, metabolic, and inflammatory pathways. In addition, obesity is strongly linked to reproductive function or organ pathologies, including endometrial hyperplasia, endometrial carcinoma^6^, and polyendocrine metabolic ovarian syndrome (PMOS). PMOS is the most common endocrine disorder in this population and up to 90% of PMOS patients are affected by overweight or obesity^7^. While both obesity and PMOS are known to impair ovarian function, their contribution to endometrial dysfunction and implantation failure remain under-investigated. GLP-1RAs, such as semaglutide (SE), have gained clinical prominence for the treatment of obesity and PMOS^8^. Between 2020 and 2023, dispensed prescriptions of GLP-1RAs among individuals aged 12 to 25 years increased by over 590% in US population^9^. Notably, among young adults (aged 18 to 25 years), women accounted for approximately 75% of all prescriptions, highlighting a substantial rise in use among females of reproductive age^9^. Currently, due to paucity of evidence in humans, the recommendations advise against the use of GLP-1RA within two months prior to planned conception, as a washout period is required due to potential, though unconfirmed, risks to the fetus^10^. Animal studies have raised concerns about the potential teratogenicity of GLP-1Ras^11^. However, human evidence remains inconclusive. Recent observational cohorts in women with type 2 diabetes or obesity did not show increased risks of major congenital malformations with early pregnancy exposure compared to the usage of insulin^12,13^. Extending beyond fetal safety, in a cohort of nearly 4,300 women who received GLP-1RA therapy within two years prior to conception, exposure was associated with a lower risk of developing gestational diabetes mellitus and hypertensive disorders of pregnancy^14^. Emerging evidence, including reports of unplanned pregnancies during GLP-1RA treatment, suggests that GLP-1RA therapy may enhance endometrial receptivity and implantation^11^, likely due to metabolic improvements and weight loss rather than reduced contraceptive efficacy^15,16^. However, the effects of GLP-1RA therapy on the endometrium are poorly understood. Liraglutide, a GLP-1RA, has recently been shown to reduce endometrial inflammation and collagen fibrosis in preclinical models, revealing a promising new avenue for GLP-1RA–based therapies in gynecological disorders^17^. However, systematic studies investigating the uterine microenvironment and embryo implantation processes under GLP-1RA treatment are still lacking. Given the widespread use of this treatment in women of reproductive age, this knowledge gap carries important clinical and safety implications. Here, we sought to determine the effects of SE on key determinants of reproductive success, including endometrial receptivity, stromal decidualization, early embryo development, and implantation. Thus, we employed advanced human in vitro models, combining paired endometrial epithelial organoids (EEOs) and stromal cells (ESCs) from healthy fertile donors with human embryonic stem cell derived embryo models, blastoids, to interrogate early developmental and implantation processes. By integrating these complementary systems, our study provides the first mechanistic insights into how SE may influence human reproduction. An overview of the experimental design is provided in **Fig. 1**.

**Fig. 1.**
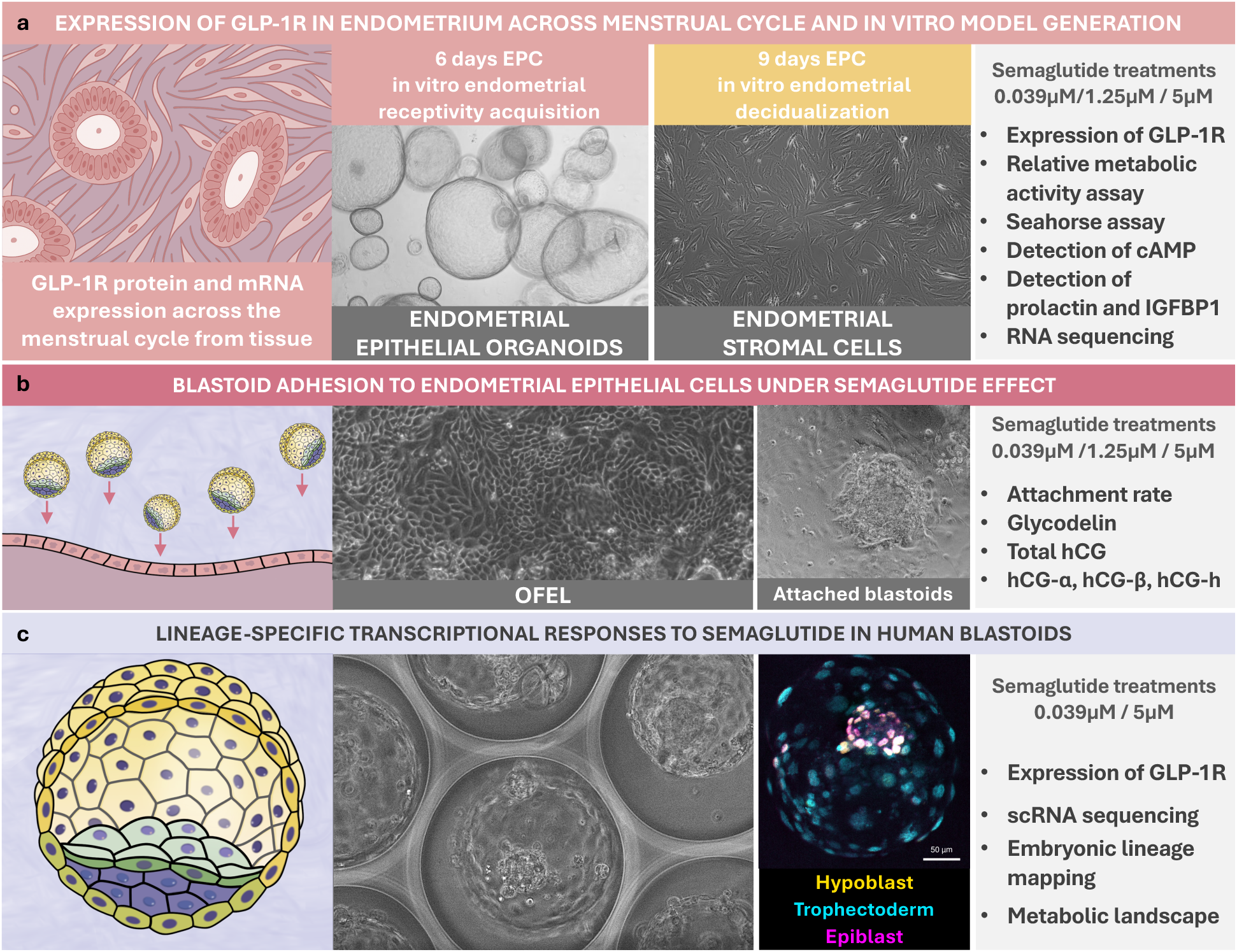
Schematic overview of the study design. **a,** GLP-1R expression was examined across the menstrual cycle in human endometrial tissue and in the generated *in vitro* endometrial models. Endometrial epithelial organoids were exposed to estrogen, progesterone, and the stable cAMP analogue 8-bromoadenosine 3′,5′-cyclic monophosphate (8-Br-cAMP; together, EPC) to mimic the receptive endometrium, while paired primary stromal cells were induced to decidualize under the same EPC stimulation. Paired in vitro endometrial models were subsequently treated with semaglutide (0.039, 1.25, and 5 μM) and assessed for several readouts including GLP-1R expression, metabolic activity, bioenergetic profiling (Seahorse), cAMP production, prolactin and IGFBP1 secretion, and transcriptomic profiling by RNA sequencing. **b,** To model embryo– endometrium interactions, blastoids were co-cultured with an *in vitro* endometrial epithelial layer (OFEL) under semaglutide treatment (0.039, 1.25 and 5 μM). Attachment rate, glycodelin secretion, and human chorionic gonadotropin (hCG) isoforms (hCG, hCG-α, hCG-β, and hyperglycosylated hCG, hCG-h) were quantified to evaluate implantation-relevant functional outputs. **c,** Human blastoids were exposed to semaglutide (0.039 and 5 μM) to define lineage-specific responses. GLP-1R expression was assessed by qPCR and immunofluorescence, while single-cell RNA sequencing enabled embryonic lineage mapping and characterization of transcriptional and metabolic programs promoted by semaglutide.

## Results

### GLP1-R expression in the human endometrium

We assessed GLP-1R expression in endometrial tissue across menstrual cycle phases by immunohistochemistry (IHC) and quantified mRNA levels by qPCR. We demonstrated that GLP-1R protein expression in the endometrium is menstrual cycle-dependent and predominantly localized in the glandular and luminal epithelium, with undetectable expression in the stroma (**Fig. 2a–d**). The expression was absent or minimal in the proliferative phase (**Fig. 2a**), however increased in the secretory phase, including early, mid, and late secretory phases of the menstrual cycle (**Fig. 2b-d**). Quantitative analysis of the GLP-1R positive epithelial area fraction (%) revealed significantly higher expression in the secretory compared with the proliferative phase (P < 0.0001, Kruskal-Wallis test), peaking in the mid-secretory phase, where the proportion of glandular epithelium positive for GLP1R was the highest. Pairwise comparisons confirmed significantly greater expression in the mid-secretory endometrium compared with both proliferative (p < 0.0001, Kruskal-Wallis test) and early secretory phases (p < 0.001, Kruskal-Wallis test), and to the late secretory endometrium (p < 0.05, Kruskal-Wallis test) (**Fig. 2e**). *GLP-1R* mRNA was detectable at low levels in endometrium throughout the menstrual cycle, with no significant variation between phases (**Extended Data Fig. 1a**). We next assessed GLP-1R expressions at both transcript and protein levels in primary EEOs and ESCs. In EEOs, GLP-1R protein and mRNA were detectable (**Fig. 2f, Extended Data Fig. 1b**), with no differences in stimulation with estradiol (E2) compared to E2, progesterone (P4) and 8-bromo-cAMP (EPC). However, expression in ESCs was below the detection threshold.

**Fig. 2.**
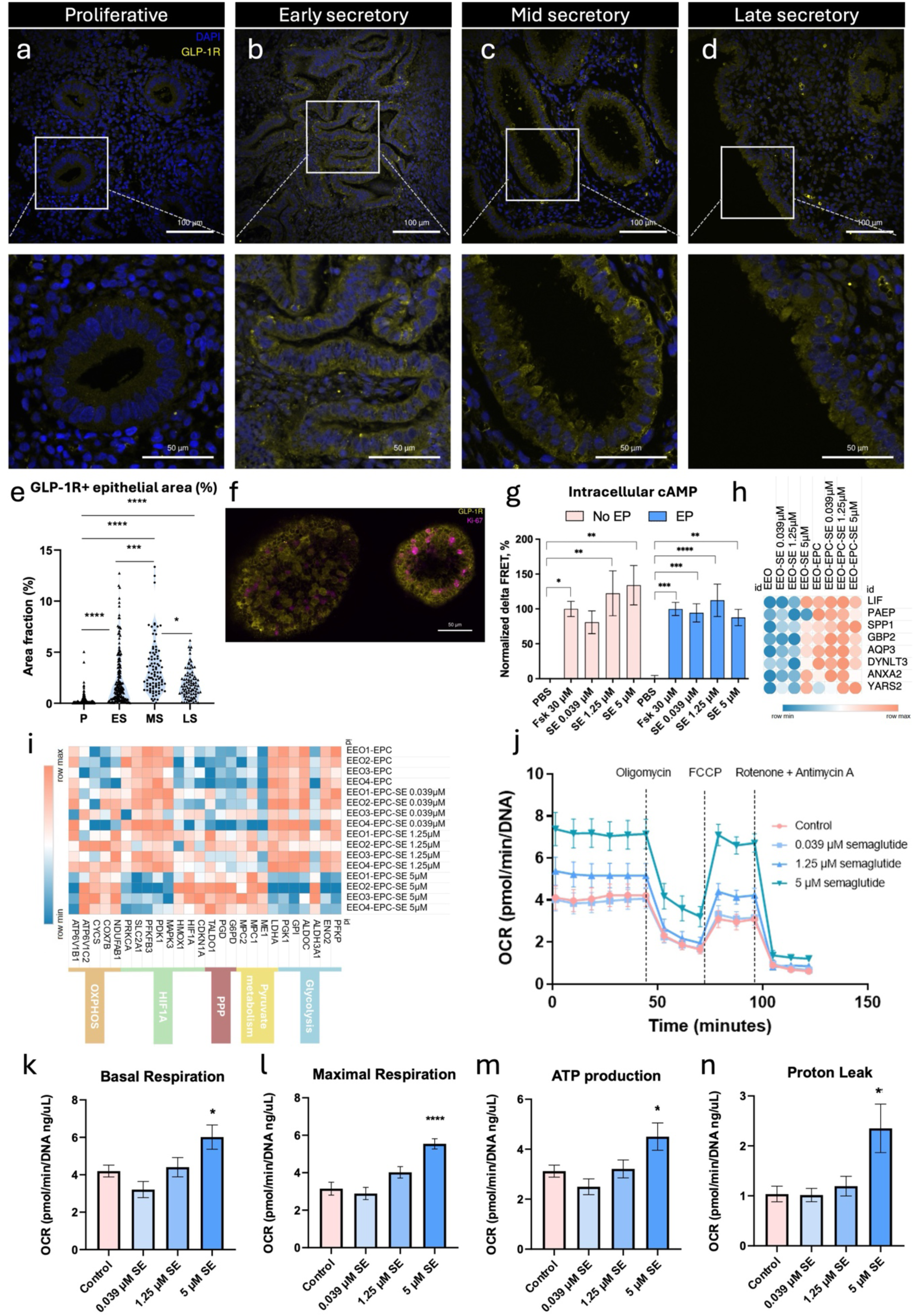
GLP-1R expression and functional responses to semaglutide (SE) in endometrial tissue biopsy and endometrial epithelial organoids (EEOs) from healthy fertile women. **a–d**, Representative images of immunohistochemical staining of GLP-1R (yellow) in endometrial tissue from healthy fertile women during the proliferative (P, N=5) (**a**), early secretory (ES, N=5) (**b**), mid-secretory (MS, N=5) (**c**), and late secretory (LS, N=5) (**d**) phases of the menstrual cycle. Nuclei were stained with DAPI (blue). Scale bar, 100 µm and magnification 50 µm. **e**, Quantification of the GLP-1R positive glandular epithelial area fractions (%) for the P, ES, MS and LS phases of the endometrial tissue. Each violin represents distribution, median, and range of the feature for each group. Asterisks indicate statistically significant differences between the groups. Statistical significance was determined using a Kruskal-Wallis test followed by Dunn’s multiple comparisons test (*p < 0.05, ***p < 0.001, ****p < 0.0001). **f,** Representative immunofluorescence image of EEOs stained for GLP-1R (yellow) and Ki-67 (pink). Scale bar, 50 µm. **g**, Normalized ΔFRET% in endometrial epithelial cells treated with forskolin (Fsk; positive control) or semaglutide (SE) (0.039, 1.25, or 5 μM) in the absence or presence of estrogen and progesterone (EP). Error bars represent the Standard Error of the Mean (SEM) calculated from two independent biological replicates, with each experiment performed in technical triplicates. Statistical significance was determined using a one-way ANOVA followed by Dunnett’s multiple comparisons test (*p < 0.05, **p < 0.01, ***p < 0.001, ****p < 0.0001). **h**, Median expression obtained from 4 biological replicates of EEOs for endometrial receptivity genes *LIF, PAEP, SPP1, GBP2, AQP3, DYNLT3, ANXA2 and YARS2* in EEOs untreated with SE (EEO), and treated with SE (EEO-SE 0.039 μM, 1.25 μM, or 5 μM) in absence of estrogen, progesterone and cAMP (EPC). Alongside expression was assessed in EPC stimulated EEOs without SE exposure (EEO-EPC) and with SE exposure (EEO-EPC-SE 0.039 μM, 1.25 μM, or 5 μM) (log₂-transformed normalized counts). **i**, Heatmap of genes associated with metabolic pathways in EPC stimulated EEOs following SE treatment at 0.039 μM, 1.25 μM, or 5 μM (log₂-transformed normalized counts). **j–n**, Oxygen consumption rate (OCR) measured by Seahorse XF Mito Stress Test in three biological replicates of EEOs treated with SE at 0.039 μM, 1.25 μM, or 5 μM, and SE untreated EEOs as control. Data represent mean ± SEM of three biological replicates, each measured in technical triplicates, with values normalized to DNA content. **j**, Pooled and normalized OCR profiles of EEOs treated with SE. Sequential injections of oligomycin, FCCP, and rotenone/antimycin A were used to assess mitochondrial function parameters. **k–n**, Quantification of OCR-derived parameters: **k**, basal respiration; **l**, maximal respiration; **m**, ATP production and **n**, proton leak. Error bars represent the SEM. Statistical analysis was performed using a one-way ANOVA followed by Dunnett’s multiple comparisons test (*p < 0.05, **p < 0.01, ****p < 0.0001).

### Semaglutide promotes canonical GLP-1R signaling and metabolic reprogramming in endometrial epithelium without compromising receptivity

To assess SE-induced cAMP signalling, EEOs were stimulated with E2 and P4 (EP) for 6 days, while the exogenous 8-Br-cAMP component of the EPC cocktail was omitted to avoid confounding the FRET-based cAMP biosensor readout. cAMP responses were subsequently evaluated following exposure to forskolin (30 µM) or SE (0.039, 1.25 or 5 µM), alongside EEOs maintained without EP stimulation or SE. SE elicited a rapid increase in intracellular cAMP levels in case of both EP stimulated and unstimulated EEOs with respect to PBS-treated EEOs (**Fig. 2g**). Further, to recapitulate the receptive endometrial state, EEOs were stimulated with EPC for a total of six days with and without SE treatment, followed by transcriptional profiling using RNA-seq. EPC stimulation enhanced epithelial receptivity in EEOs compared to non-EPC stimulated EEOs, as confirmed by significant differential expression of 29 endometrial receptivity-associated genes, including key markers such as *PAEP* and *LIF* (**EEO-EPC vs EEO, Extended Data Table 1**). Further, the hormonal stimulation (EPC) and exposure of varying concentration of SE also showed significant differential expression of endometrial receptivity-associated genes when compared to EEO controls (EPC-SE untreated). In total, 36 (EEO-EPC-SE0.039 μM vs control), 32 (EEO-EPC-SE1.25 μM vs control) and 25 (EEO-EPC-SE5 μM vs control) genes were differentially expressed (**Extended Data Table 1**). Interestingly, the expression of key biomarkers of endometrial receptivity remained inducible with presence of SE (0.039 μM and 1.25 μM) with EPC treatment, indicating that SE, at these concentrations, does not impair receptivity (**Fig. 2h**). However, at the highest SE concentration (5 μM), *PAEP* induction was attenuated, suggesting that SE may modulate specific components of the endometrial receptivity program (**Fig. 2h, Extended Data Table 1**). Noteworthy, non-EPC stimulated EEOs exposed to 5 μM SE (EEO-SE5 μM) exhibited significantly increased expression of multiple receptivity-associated genes, including *LIF*, *GBP2*, *AQP3*, *ANXA2*, and *YARS2* compared to SE-EPC-untreated EEOs (**Fig. 2h**, **Extended Data Table 1**). These findings suggest that 5 μM SE enhances expression of receptivity genes in the endometrium even in the absence of hormonal priming.

From a metabolic perspective, NAD(P)H-dependent resazurin assay revealed a bell-shaped EEO response to SE, with enhanced relative metabolic activity at low concentrations but significant suppression at 5 µM, without compromising the cell viability (**Extended Data Fig. 1c–d**). Transcriptomic profiling of EPC-stimulated EEOs treated with 0.039 µM and 1.25 µM SE, compared to SE-untreated EPC-EEO revealed 2 downregulated differentially expressed genes (DEGs) (*USF3* and *CD99*) and no DEGs, respectively. SE treatment with 5 µM revealed coordinated downregulation of glycolytic genes (*LDHA*, *ALDH3A1*) and upregulation of genes involved in NADH/NADPH generation (*G6PD*, *PGD*, *ME1*, *TALDO1*), pyruvate metabolism (*MPC1/2*), and oxidative phosphorylation (**Fig. 2i, Extended Data Table 2**). Expression of *HIF1A* was modestly increased, whereas *SLC2A1* and MAPK pathway components (*MAPK3*, *PRKCA*) were suppressed. Gene ontology analysis highlighted downregulation of hypoxia and monosaccharide metabolic pathways, alongside increased NAD(P)(H)-related metabolic processes (**Extended Data Fig. 1e–f**). To validate metabolic dynamics in EEOs, we measured oxygen consumption rate (OCR) in three biological replicates using the Seahorse Mito Stress Test (**Fig. 2j**), which revealed a concentration-dependent increase in OCR with escalating SE exposure. It revealed that SE treatment (0.039 µM and 1.25 µM) did not alter the mitochondrial functions, whereas treatment with 5 μM SE significantly increased basal and maximal respiration (P < 0.05, one-way ANOVA; P < 0.0001, one-way ANOVA, respectively), ATP production (P < 0.05, one-way ANOVA), and proton leak (P < 0.01, one-way ANOVA) in EEOs compared to control (SE-untreated EEOs) (**Fig. 2k–n**), indicative of elevated mitochondrial oxidative metabolism. Importantly, basal and maximal respiration (**Fig. 2k-l**), ATP production (**Fig. 2m**) and proton leak (**Fig. 2n**) remained within physiological limits, consistent with preserved mitochondrial integrity and suggesting increased mitochondrial content rather than mitochondrial stress. Together, these findings demonstrate that SE promotes canonical GLP-1R signaling and drives a metabolic shift towards oxidative phosphorylation in endometrial epithelium, where GLP-1R expression peaks during the mid-secretory phase and SE exposure does not detrimentally affect key molecular markers of receptivity.

### Semaglutide exhibited an anti-proliferative effect and impaired endometrial stromal decidualization

Similar to EEOs, ESCs were assessed for SE-induced changes in intracellular cAMP and metabolic activity. For cAMP measurements, ESCs were stimulated with E2 and P4 only, omitting exogenous 8-Br-cAMP to avoid interference with the biosensor readout, whereas metabolic activity was evaluated by resazurin reduction. In parallel, ESCs were stimulated with the full EPC cocktail for 9 days to induce decidualization, followed by transcriptomic profiling using RNA sequencing. In contrast to EEOs, in which SE increased intracellular cAMP levels, cAMP responses in SE-treated ESCs remained low relative to the forskolin control, with a significant increase observed only at 5 µM SE (**Fig. 3a**). Using the resazurin-resorufin assay, we found that SE significantly suppressed metabolic activity in ESCs at concentrations of 1.25 μM (P < 0.0001, one-way ANOVA) and 5 μM (P < 0.0001, one-way ANOVA) without compromising cell viability, whereas exposure to 0.039 μM had no detectable effect (**Extended Data Fig. 2a, Extended Data Fig. 2b**). During EPC-induced in vitro decidualization, ESCs secreted human insulin-like growth factor binding protein 1 (IGFBP-1) and prolactin (PRL), consistent with successful decidual transformation (**Fig. 3b-c**). SE exposure significantly decreased IGFBP-1 secretion at 1.25 μM (P < 0.05, one-way ANOVA) and 5 μM (P < 0.0001, one-way ANOVA) and significantly reduced PRL secretion in a concentration-dependent manner (0.039 μM P < 0.01; 1.25 μM P < 0.001; 5 μM P < 0.0001, one-way ANOVA) (**Fig. 3b-c**), highlighting the inhibitory effect of SE on ESC decidualization. Transcriptomic analysis of EPC-stimulated and SE-treated ESCs confirmed concentration-dependent decreased expression of *PRL* and *IGFBP-1*, consistent with the corresponding changes in protein secretion (**Fig. 3d**, **Extended Data Fig. 2c**). EPC treated ESCs with different concentration of SE compared to control (EPC-ESCs untreated with SE) revealed that low concentration of SE (0.039 μM) induced no significant gene expression changes, while 1.25 μM downregulated only three genes, including *ITGA10, THBD* and *SKA3*, suggesting early suppression of mitotic activity. In contrast, treatment with 5 μM SE led to the significant downregulation of 120 genes and upregulation of 12 genes, with enrichment analysis indicating suppression of cell cycle progression (**Extended Data Fig. 2d**), particularly at the G2/M transition (**Fig. 3e, Extended Data Table 3**). Notably, there was no differential expression of pro-apoptotic or senescence-associated genes. At 5 μM SE concentration, upregulation of genes involved in endoplasmic reticulum-associated degradation (ERAD), IRE1-XBP1s signaling, and N-glycosylation pathways were observed (**Fig. 3f, Extended Data Fig. 2e**). To determine whether these transcriptional effects were specific to decidualization, we compared SE-induced gene-expression changes at 5 µM in decidualizing (EPC) and estrogen-stimulated (E2) ESCs. The fold changes of SE-responsive genes were strongly correlated between the two hormonal contexts (Pearson’s r = 0.91), indicating a highly concordant transcriptional response. These findings suggest that the anti-proliferative effect of SE represents a broader response and is not restricted to hormonal stimulation in ESCs (**Fig. 3g**).

**Fig. 3.**
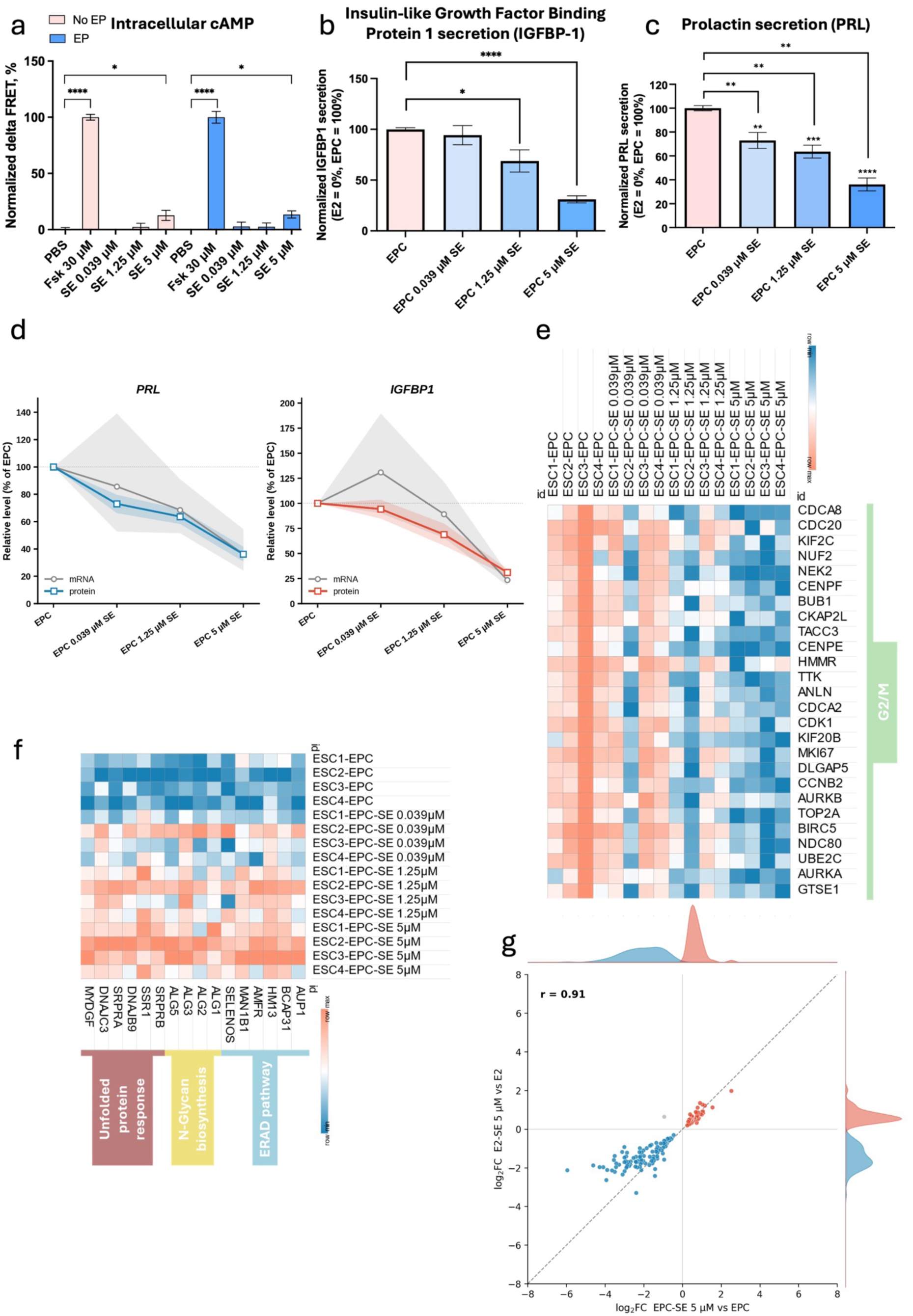
Semaglutide (SE) alters decidualization and cell-cycle programs in human endometrial stromal cells (ESCs). **a**, Normalized change in Förster resonance energy transfer efficiency (ΔFRET%) reporting intracellular cAMP levels in ESCs treated with forskolin (Fsk) or SE (0.039 μM, 1.25 μM, or 5 μM), with or without estrogen and progesterone (EP/ NO EP). **b**, Insulin-like Growth Factor Binding Protein 1 secretion (IGFBP-1) secretion measured by ELISA after 9 days of EPC stimulation in SE-treated (EPC 0.039 μM, 1.25 μM, or 5 μM SE) and SE-untreated (ESC-EPC) ESCs. Error bars represent the SEM calculated from four independent biological replicates, with each experiment performed in technical duplicates. Statistical significance was determined using a one-way ANOVA followed by Dunnett’s multiple comparisons test (***p < 0.001, ****p < 0.0001). **c**, Prolactin (PRL) secretion measured by ELISA after 9 days of EPC stimulation in SE-treated (EPC 0.039 μM, 1.25 μM, or 5 μM SE) and SE-untreated (ESC-EPC) ESCs. Error bars represent the Standard Error of the Mean (SEM) calculated from three independent biological replicates, with each experiment performed in technical duplicates. Statistical significance was determined using a one-way ANOVA followed by Dunnett’s multiple comparisons test (*p < 0.05, **p < 0.01, ***p < 0.001, ****p < 0.0001). **d**, Dose-dependent changes in PRL and IGFBP-1 at the protein and transcript level in EPC-stimulated ESCs treated with SE (0.039 μM, 1.25 μM, or 5 μM), expressed as a percentage of the SE-untreated EPC control (100%). Grey lines show transcript levels (RNA-seq normalized counts; geometric mean of donor-paired ratios) and coloured lines show secreted protein (ELISA; PRL, blue; IGFBP-1, red). Points represent means and shaded bands the SEM (transcript, n = 4 donors; protein, n = 3 (PRL) and n = 4 (IGFBP-1) independent biological replicates). **e**, Heatmap of G2/M phase-related genes in ESCs treated with EPC and SE (EPC-SE-0.039 μM, 1.25 μM, or 5 μM) (log₂-transformed normalized counts). **f**, Heatmap of genes associated with the unfolded protein response, N-glycan biosynthesis, and ER-associated degradation (ERAD) pathways in ESCs treated with EPC and SE (EPC-SE-0.039 μM, 1.25 μM, or 5 μM) (log₂-transformed normalized counts). **g**, Concordance of semaglutide-induced transcriptional changes between decidualizing and estrogen-only ESCs. Each point represents a semaglutide-responsive gene (n = 145 genes detected in both comparisons), plotted by its log₂ fold change in decidualizing cells (EPC-SE 5 µM vs EPC; x-axis) against that in estrogen-only cells (E2-SE 5 µM vs E2; y-axis). Genes down-regulated in both conditions are shown in blue and genes up-regulated in both in red; the dashed line denotes the identity (y = x) and marginal density plots show the distribution of fold changes along each axis.

### Semaglutide enhances blastoid adhesion to endometrial cells under hormone-free conditions

To investigate the effects of SE on embryo–endometrial interactions, we employed an in vitro implantation model to assess human blastoid attachment to endometrial epithelial cells referred to as open-faced endometrial layer (OFEL). Human blastoids were generated from naïve H9 human embryonic stem cells (ESCs) and transferred to hormonally stimulated OFELs under the effect of SE. Hormonal priming promoted blastoid adhesion, expansion, and interaction with the OFEL (**Fig. 4a, 4b**). In contrast, hormone deprivation substantially impaired blastoid attachment to the OFEL (**Fig. 4b**). When blastoids were transferred onto OFEL treated with SE, all concentrations significantly increased blastoid adhesion under hormone-free conditions to levels comparable with hormonally primed controls (0.039 µM, P < 0.0001; 1.25 µM, P < 0.001; 5 µM, P < 0.001; two-ways ANOVA when compared to No EPCX and SE) (**Fig. 4b**). Under hormonally stimulated conditions, SE treatment did not further enhance or disturb blastoid attachment. Additionally, we assessed the concentrations of secreted proteins serving as biomarkers of endometrial receptivity (*PAEP*-encoding glycodelin) or trophoblast differentiation/early embryonic signaling (human chorionic gonadotropin, hCG). Glycodelin secretion was strictly dependent on hormonal stimulation (EPCX) and was undetectable in its absence (**Extended Data Fig. 3a**). Under blastoid attachment with hormonal stimulation, 0.039 and 1.25 µM SE significantly increased glycodelin secretion relative to OFEL without SE (P < 0.05, two-way ANOVA; **Extended Data Fig. 3a**). By contrast, 5 µM SE markedly reduced glycodelin secretion relative to OFEL without SE and to OFEL exposed to 0.039 or 1.25 µM SE, irrespective of blastoid presence (**Fig. 4c**). This reduction is consistent with transcriptomic data showing that 5 μM SE decreases expression of the glycodelin-encoding gene *PAEP* (**Fig. 2h**).

**Fig. 4.**
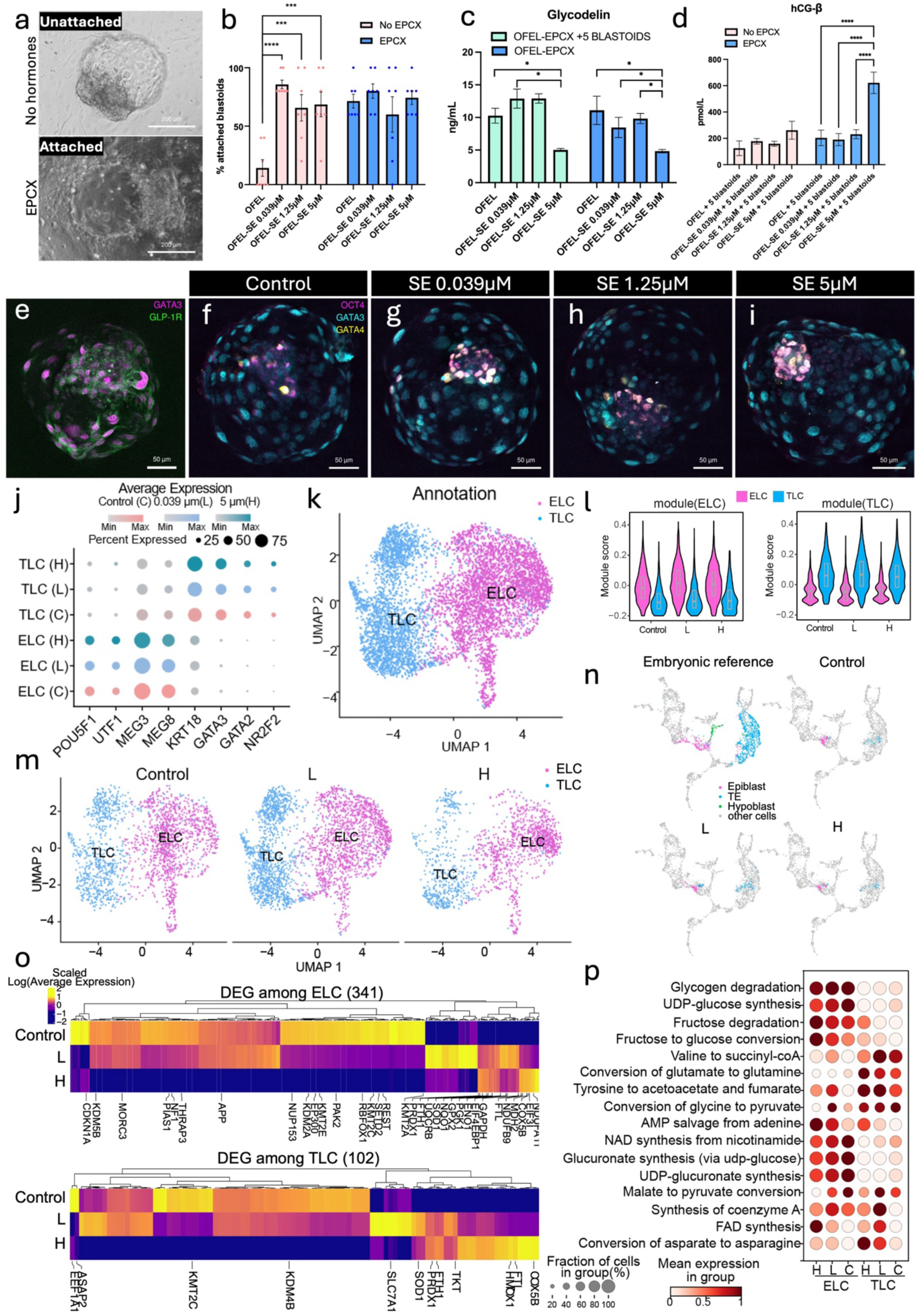
Effects of semaglutide (SE) on blastoid attachment, differentiation, and lineage-specific responses. **a**, Representative bright-field images of non-attached blastoids co-cultured with endometrial epithelial cells in the absence of estrogen, progesterone, cAMP, and XAV-939 (EPCX), and attached blastoids under EPCX conditions. Scale bar, 200 µm. **b**, Percentage of blastoids attached to the open-faced endometrial layer (OFEL) following treatment with SE (OFEL-SE-0.039 μM, 1.25 μM, or 5 μM) alone or in combination with EPCX. Error bars represent the Standard Error of the Mean (SEM) calculated from two biological replicates, with two independent experiments (N=4). Each experiment was performed in five technical replicates. Statistical significance was determined using a two-way ANOVA followed by Šidák’s multiple comparisons test (***p < 0.001, ****p < 0.0001); SE significantly increased attachment relative to no-EPCX, SE-untreated controls, whereas under EPCX-primed conditions SE did not significantly alter attachment beyond the level already achieved by hormonal priming alone. **c**, Glycodelin levels measured by time-resolved fluorescence (TRF) immunoassay in spent culture media from OFEL exposed to EPCX conditions, and with five blastoids per well or without blastoids, treated with SE (OFEL-SE-0.039 μM, 1.25 μM, or 5 μM). **d**, hCG-β secretion measured by TRF immunoassay in OFEL cultures treated with SE containing five blastoids per well (OFEL-SE-0.039 μM, 1.25 μM, or 5 μM + 5 blastoids), either without or with EPCX. Error bars represent the SEM calculated from two biological replicates, with two independent experiments (N=4). Each experiment was performed in five technical replicates. Statistical significance was determined using a two-way ANOVA followed by Tukey’s multiple comparisons test (*p < 0.05, ****p < 0.0001); comparisons are made against the corresponding SE-untreated condition within the same hormonal status (with or without EPCX). **e–i**, Representative immunofluorescence images of blastoids: **e**, GLP-1R expression (green) co-stained with the trophectoderm marker GATA3 (purple); **f–i**, blastoids stained for OCT4 (purple), GATA3 (cyan), and GATA4 (yellow) for SE-untreated (**f**) and SE-treated (0.039 μM (**g**), 1.25 μM (**h**), 5 μM (**i**)) conditions. Scale bar, 50 µm. **j**, Dot plot showing average gene expression per cell for epiblast-like cells (ELC) and trophectoderm-like cells (TLC) in control (C) and SE-treated blastoids at 0.039 μM (L) and 5 μM (H). **k**, Uniform manifold approximation projection (UMAP) of the annotated TLC and ELC lineages. **l**, Module scores for ELC and TLC gene sets in blastoids treated with SE 0.039 μM (L) and 5 μM (H) or SE-untreated control. **m**, UMAP projection of single-cell RNA-seq data from control and SE-treated blastoids (L or H), with cells annotated as ELCs and TLCs. **n,** UMAP projection of single-cell RNA-seq data from SE-treated blastoids 0.039 μM (L) and 5 μM (H) and SE-untreated blastoids (control) mapped onto a human embryo single-cell reference atlas. **o**, Lineage-specific transcriptomic responses (log average expression) to SE: heatmaps of consistently differentially expressed genes (DEGs) in ELCs compared to SE-untreated blastoids (control) and TLC compared to control showing upregulation of glycolysis, oxidative phosphorylation, and NRF2 pathway genes, with concurrent downregulation of histone-modifying enzymes following SE treatment; 0.039 μM (L) and 5 μM (H). **p,** Dot plot showing the relative enrichment and mean expression levels of genes involved in key metabolic pathways across the identified cell clusters (ELC and TLC) in SE-treated blastoids 0.039 μM (L) and 5 μM (H) and SE-untreated blastoids (controls). The size of each dot indicates the fraction of cells within each cluster expressing genes associated with the given pathway, while the color intensity represents the mean expression level.

Furthermore, hormonal stimulation significantly increased secretion of intact hCG (P < 0.001; two-way ANOVA) and hyperglycosylated hCG (hCG-h; P < 0.01; two-way ANOVA) compared with unstimulated conditions in the attachment assay, whereas secretion of the free α (hCG-α) (**Extended Data Fig. 3b–d**) and β (hCG-β) subunits remained unchanged (**Fig. 4d**). Intact hCG secretion was elevated by hormonal stimulation but was not further increased by SE at any concentration, indicating that intact hCG production was driven by hormonal priming rather than by SE (**Extended Data Fig. 3b**). In contrast, 5 µM SE selectively increased free hCG-β secretion compared with the corresponding hormonally stimulated SE-untreated control (P < 0.0001; two-way ANOVA) (**Fig. 4d**) without affecting hCG-α (**Extended Data Fig. 3d**). In addition, under 5 µM SE treatment, hCG-h secretion was significantly higher in hormonally stimulated than in unstimulated conditions (P < 0.05; two-way ANOVA; **Extended Data Fig. 3c**), indicating a shift toward a hyperglycosylated hCG-dominant state. Collectively, these findings indicate that intact and hCG-h secretion were driven primarily by hormonal priming, whereas semaglutide acted selectively at the highest concentration (5 µM) to increase the free hCG-β subunit and hCG-h without raising intact hCG or the α subunit.

### Lineage-specific transcriptional responses to semaglutide reveal metabolic and epigenetic reprogramming in human blastoids

To assess the effect of SE on blastoid formation and lineage development, SE-treated and control blastoids were collected on day 6. Transcriptional lineage-specific responses were analyzed by single-cell RNA sequencing (scRNA-seq), and GLP-1R expression was evaluated by immunofluorescence (IF) and qPCR. Blastoids expressed GLP-1R at both the protein (**Fig. 4e**) and mRNA levels (**Extended Data Fig. 4a**), and SE treatment did not significantly alter *GLP-1R* transcript abundance. IF confirmed lineage-specific markers for epiblast (OCT4), trophectoderm (GATA3), and hypoblast (GATA4) in untreated blastoids (**Fig. 4f**). SE treatment with 0.039 μM, 1.25 μM and 5 μM revealed the presence of all three cell lineages (**Fig. 4g-i**). Clustering by cell-specific markers following scRNA-seq identified an epiblast-like cell (ELC) population characterized by *POU5F1*, *UTF1*, *MEG3* and *MEG8* expression, and a trophectoderm-like cell (TLC) population marked by *GATA2*, *GATA3*, *NR2F2* and *KRT18* expression in low (L) 0.039 μM and high (H) 5 μM SE conditions (**Fig. 4j**). UMAP projection of blastoids from all integrated conditions further resolved two transcriptionally distinct populations corresponding to ELCs and TLCs (**Fig. 4k**). To determine whether SE treatment affected lineage identity, we compared epiblast- and trophectoderm-associated module scores across ELCs and TLCs under all conditions. Epiblast-associated module scores were uniformly elevated in ELCs, whereas trophectoderm-associated module scores were enriched in TLCs (**Fig. 4l**). These patterns were maintained regardless of SE treatment, indicating that lineage identity remained stable. Further, UMAP projection of blastoids treated with 0.039 μM and 5 μM SE revealed broadly similar patterns of cell-type distribution to those observed in controls (SE-untreated blastoids) (**Fig. 4m**). Consistent with the limitations of current blastoid models, hypoblast cells were sparse in the scRNA-seq post-quality control dataset based on the expression of hypoblast-specific markers and unable to form distinct cluster (**Extended Data Fig. 4b**), although GATA4 IF confirmed their presence (**Fig. 4f**). Integration with a reference single-cell atlas of human embryos^18^ confirmed ELC and TLC identities and indicated that the blastoids recapitulate the human blastocyst epiblast and trophectoderm lineages (**Fig. 4n**). Lineage-specific transcriptional responses to SE were defined by identifying DEGs consistently altered at both 0.039 µM (L) and 5 µM (H) relative to their respective controls. In the ELC, we observed upregulation of genes involved in glycolysis (*GAPDH, MDH2, ENO1, PGK1*), oxidative phosphorylation (*UQCRB, COX5B, NDUFB10*), and the NRF2 pathway (*FTL, FTH1, PRDX1, HSP90AA1*) (**Fig. 4o, Extended Data Fig. 4c**). The downregulated genes were involved in histone modification, particularly histone demethylases (*KDM5B, KDM6A, KDM7A, KDM2A*) (**Fig. 4o, Extended Data Fig. 4c**). These findings suggest that SE enhances glycolytic and mitochondrial activity, potentially activating the NRF2 pathway as a protective response to reactive oxygen species accumulation. In the TLC we observed similar responses as genes involved in oxidative phosphorylation (*UQCRB, COX5B, NDUFB9*), and the NRF2 pathway (*FTL, FTH1, PRDX1, NQO1*) were upregulated, while genes encoding histone-modifying enzymes such as demethylase (*KDM4B*) and methyltransferase (*KMT2C*) were downregulated (**Fig. 4o, Extended Data Fig. 4c**), highlighting that the TLC and ELC respond similarly to the GLP-1RA. To further understand the metabolic changes in the SE-treated blastoids we performed a single-cell metabolic analysis that predicts the pathway-specific metabolic activity^19^. This analysis revealed lineage-specific differences between the ELC and TLC in the control (untreated) blastoids. Glycolysis-associated carbohydrate metabolism, including glycogen degradation, UDP-glucose synthesis, fructose degradation, and fructose-to-glucose conversion, were more active in the ELC, whereas reactions linked to mitochondrial oxidative metabolism and feeding into the tricarboxylic acid (TCA) cycle such as valine-to-succinyl-CoA, glutamate-to-glutamine, tyrosine-to-acetoacetate/fumarate, and glycine-to-pyruvate conversion were upregulated in TLC (**Fig. 4p, Extended Data Table 4**). Furthermore, SE treatment induced transcriptional metabolic reprogramming in both the TLC and ELC, consistent with a shift towards oxidative metabolism. In the TLC, AMP salvage and malate-to-pyruvate conversion were increased, while NAD synthesis and UDP-glucose–related pathways were reduced, indicating enhanced mitochondrial activity. In the ELC, upregulation of coenzyme A and FAD synthesis supported sustained oxidative phosphorylation, accompanied by adaptive changes in aspartate and nucleotide metabolism. Collectively, these results suggest that SE promotes mitochondrial efficiency and redox adaptation, while remodeling metabolic pathways that intersect with epigenetic regulation, combined with the reduced expression of histone-modifying enzymes. These data indicate coordinated metabolic and transcriptional reprogramming, although direct epigenetic changes remain to be established (**Fig. 4p, Extended Data Table 4**).

## Discussion

SE, a GLP-1RA, shows potential in improving female fertility by modulating metabolic imbalance and overweight. Traditionally, weight loss has been thought to restore ovulation by correcting hormonal imbalances. However, the effects of SE on the endometrium remain poorly understood. It has been previously reported that GLP-1R expression in endometrial cancer was correlated with higher survival rate, suggesting a better prognosis^20^. However, data regarding its expression patterns and dynamics in non-pathological tissues, as well as its menstrual cycle–dependent regulation, remained unavailable. Our findings reveal a previously unrecognized role for GLP-1R in the healthy human endometrium, characterized by dynamic, cycle-dependent expression predominantly within glandular and luminal epithelial cells during the secretory phase, with no detectable expression in stromal cells. The predominance observed during the secretory phase, when embryo implantation is expected, underscores the need to elucidate its potential role in regulating embryo implantation and early embryonic development. The absence of detectable GLP-1R protein in stromal cells, despite low but consistent GLP-1R mRNA levels in whole endometrial tissue, underscores epithelial enrichment as the principal source of GLP-1R activity in vivo. This was further substantiated by the robust detection of mRNA and protein expression of GLP-1R in EEOs, coupled with its absence in primary ESCs. Importantly, the observed discrepancy between stable GLP-1R mRNA levels and dynamic protein expression across the menstrual cycle suggests that receptor abundance is regulated post-transcriptionally, likely via mechanisms such as translational control, protein stability, or receptor trafficking^21^.

Elevated cAMP is the canonical downstream response to GLP-1R activation, as the Gα proteins coupled to the receptor stimulate adenylyl cyclase to increase intracellular cAMP^11^. Consistent with this mechanism, we detected expression of GLP-1R in EEOs and observed robust cAMP elevation following ligand stimulation. This aligns with the established pathways in which elevated cAMP activates PKA and EPAC2, driving transcription of genes like *LIF*, *PTGS2* and *PGE2* that play a vital role in endometrial receptivity^11^. In our study, we show that EPC-EEOs treated with 0.039 μM and 1.25 μM SE exhibit unchanged expression of receptivity-associated genes compared to untreated EPC-EEOs, indicating that SE does not exert a deleterious impact on receptivity. However, at the higher concentration of 5 µM, endometrial receptivity was impaired to some extent, as reflected by reduced number of upregulated and increased number of downregulated receptivity-associated genes, including suppressed *GPX3*, which regulates oxidative stress by reducing reactive oxygen species to support a receptive endometrial environment^22,23^. Notably, *PAEP* expression was not significantly upregulated at the SE concentration of 5 µM, despite being a well-documented marker considered essential for successful embryo implantation^24^. These findings suggest that higher concentrations of SE may compromise endometrial receptivity. This pattern is consistent with the possibility that excessive GLP-1R stimulation induces receptor desensitization or downstream feedback inhibition, thereby attenuating the cAMP signaling axis that supports receptivity. Nonetheless, the potential contribution of off-target effects at 5 µM cannot be excluded, particularly given the likelihood of biased signaling. Notably, the receptivity-associated genes analysed were selected from literature^24,25^ describing expression profiles in human endometrial tissue, which encompasses all major cellular populations, while experimentally, we examined epithelial in vitro model. This difference in cellular composition likely accounts for the lower number of overlapping genes and certain discrepancies with tissue-based panels, including contrasting trends for genes such as *EDN3*, *MMP7* and *CRABP2,* which were upregulated in our model despite being reported as downregulated in whole-tissue analyses. Collectively, at the transcriptional level, treatment of EEOs with SE (0.039 μM) did not induce any significant changes compared with untreated controls, indicating that GLP-1R activation at this concentration is unlikely to disrupt endometrial function. Interestingly, even at the higher concentration of 1.25 μM, transcriptional profiles remained significantly unchanged, suggesting that SE does not compromise endometrial integrity across a broad concentration range. These observations provide initial reassurance that exposure to SE during the window of implantation is unlikely to adversely affect endometrial receptivity. Still, in condition without steroid hormone priming, the EEOs show gene expression/transcriptomic changes consistent with endometrial receptivity after exposure to SE at the highest concentration. Although these findings suggest that GLP-1R activation can modulate receptivity-associated pathways independently of steroid hormones, its relevance to improved implantation competence or reproductive outcomes requires further investigation.

Furthermore, our study demonstrates that SE exerts a bell-shaped effect on EEO metabolism revealed by resazurin-resorufin assay, with 0.039 μM and 1.25 μM enhancing cellular metabolic activity and 5 μM suppressing it. SE treatment with 5 μM promoted a coordinated metabolic reprogramming away from glycolysis, as indicated by downregulation of *LDHA* and *ALDH3a1*, and toward oxidative metabolism, reflected by the upregulation of key regulators of NADH/NADPH generation (*G6PD*, *PGD*, *ME1*, *TALDO1*), pyruvate transport (*MPC1/2*), and oxidative phosphorylation. Mechanistically, although *HIF1A* expression was modestly increased, consistent with activation of hypoxia-responsive pathways, glucose uptake (*SLC2A1*) and MAPK signaling were suppressed, indicating that SE selectively enhances mitochondrial metabolism without broadly stimulating proliferative or glycolytic pathways. Transcriptomic changes supporting oxidative phosphorylation have been shown to be consistent with enhanced mitochondrial function and energy production, whereas mitochondrial dysfunction particularly in insulin-resistant or high-fat diet models promotes oxidative stress and compromises endometrial receptivity^26,27^. The regulation of oxidative phosphorylation and metabolism by SE has been reported in several different tissues and pathophysiological conditions. In a mouse model of heart failure, SE preserved mitochondrial integrity and redirected cardiac metabolism toward oxidative glucose utilization via NR4a1, thereby attenuating maladaptive remodeling^28^. Supporting this mechanistic link, short-term GLP1-RA treatment increased glucose oxidation and improved cardiac energetics in a rat model of high-fat diet–induced diastolic dysfunction^29^. Consistent with its mitochondrial actions in the heart, SE also normalized mitochondrial morphology and function in a model of autosomal dominant polycystic kidney disease^30^. In contrast, in patients with metabolic dysfunction–associated steatohepatitis, long-term SE treatment (0.4 mg, 72 weeks) was associated with downregulation of oxidative phosphorylation alongside attenuation of inflammation^31^. Collectively, these studies demonstrate that SE exerts tissue and pathophysiology-specific modulation of oxidative phosphorylation, preserving mitochondrial integrity and enhancing oxidative metabolism.

Seahorse Mito Stress assay showed that the 0.039 μM and 1.25 μM SE concentrations did not significantly alter mitochondrial function, whereas treatment with 5 μM SE increased basal and maximal respiration, ATP production, and proton leak, reflecting enhanced mitochondrial oxidative metabolism. Notably, the preservation of ATP production efficiency, respiratory control ratio, and proton leak within physiological limits indicates that this metabolic shift represents increased mitochondrial content and respiratory capacity rather than mitochondrial stress or damage^32^. These results suggest that SE promotes mitochondrial function in a manner consistent with adaptive cellular remodeling rather than dysfunction.

In ESCs, SE impaired decidualization in a concentration-dependent manner, as reflected by reduced PRL and IGFBP-1 at both the protein and mRNA levels. Transcriptomic profiling exhibited genes involved in upregulation of ERAD pathway, IRE1-XBP1s signaling, and N-glycosylation pathways in response to 5 μM SE. Activation of the IRE1–XBP1s axis has previously been shown to remodel the N-glycan composition of membrane-associated and secreted glycoproteins^33^. Therefore, these findings may indicate altered proteostasis and glycoprotein processing rather than enhanced secretory competence. Whether such changes affect the glycosylation of proteins involved in embryo–endometrium communication will require direct glycomic and functional assessment. To note, glycosylation plays important roles in maternal-fetal crosstalk^34^, thus alterations in glycosylation could influence endometrial receptivity and embryo–endometrium interactions, warranting further investigation. Moreover, these SE-associated alterations may further interfere with hormonal responses, as evidenced by reduced PRL and IGFBP-1 secretion, ultimately contributing to endometrial stromal dysfunction following SE treatment. SE also suppressed mitotic activity and downregulated cell cycle checkpoints, particularly at the G2/M transition. Consistent with our findings, animal models have demonstrated that the accumulation of unfolded proteins activates the endoplasmic reticulum–associated degradation (ERAD) pathway, thereby perturbing cell-cycle progression and ultimately impairing decidualization^35^. Previous analyses have shown that decidualization is characterized by an accumulation of stromal fibroblasts in the G1 phase of the cell cycle, suggesting that a G1-arrested state is permissive for differentiation^36^. In agreement with this concept, we observed a marked downregulation of G2/M transition– associated genes, reinforcing the notion that exit from the proliferative cycle is integral to acquisition of the decidual phenotype. Notably, SE exposure further suppressed PRL expression and downregulation of G2/M cell-cycle related genes, pointing to a disruption of the delicate balance between cell-cycle and endometrial receptivity. These data converge with prior evidence linking unfolded protein responses and ERAD activation to cell-cycle perturbation and impaired decidualization^35^, highlighting proteostasis and metabolic checkpoints as critical determinants of reproductive competence. The effect of high concentration of SE on ESCs, despite undetectable GLP-1R, may suggest off-target or non-GLP-1R-mediated mechanisms involving interactions with other G protein–coupled receptors. Alternatively, these effects may arise from low level, transient, or non-canonical GLP-1R activity below detection thresholds. Although GLP-1R was not detectable in ESCs, SE treatment at higher concentrations caused a modest increase in intracellular cAMP, suggesting that the suppression of mitosis and cell proliferation may result from a direct off-target effect. Our results therefore extend current knowledge by highlighting a potential cell cycle–related mechanism through which SE may affect stromal cell physiology.

Furthermore, in our study, SE showed an effect on the in vitro blastoid attachment model. Interestingly, SE enhanced the blastoid adhesion in hormone-deprived conditions, suggesting a potential ability to compensate for the lack of endocrine support through cAMP-dependent signalling pathways. However, the endometrium is inherently hormone-dependent, with its homeostasis and cellular activity continuously modulated by cyclical fluctuations of estrogen and progesterone; thus, a hormone-free state comparable to in vitro conditions is physiologically implausible. Under hormonal stimulation, SE was shown to exert dual effects on the implantation microenvironment. Treatment with 5 µM SE markedly decreased glycodelin secretion from the OFEL, a well-established marker of endometrial receptivity and embryo implantation^24,37^. This reduction may reflect altered epithelial secretory function, indicating that SE exerts concentration-dependent effects, whereby lower concentrations (0.039 and 1.25 µM) may promote epithelial–embryo adhesion, whereas the higher concentration (5 µM) may alter the immunomodulatory milieu of the implantation microenvironment. This decreased glycodelin level was consistent with transcriptomic suppression of *PAEP* expression in EEOs by the same SE concentration suggesting coordinated regulation of glycodelin at both the transcriptional and protein levels. Beyond its role in endometrial receptivity, glycodelin exerts immunomodulatory functions by influencing cell survival, proliferation, and differentiation^45^. During pregnancy, glycodelin orchestrates immune adaptation by modulating the T helper cell balance, expanding regulatory T cells, and driving monocyte differentiation toward decidual macrophage–like phenotypes, mechanisms that collectively reinforce feto–maternal tolerance^38^. Glycodelin is therefore essential for establishing and maintaining immune homeostasis at the feto–maternal interface, safeguarding the fetus from maternal immune rejection and promoting proper placental development. Clinically, reduced circulating glycodelin A levels are associated with recurrent spontaneous abortion^39^, highlighting its pivotal role in reproductive immunology and potential as a biomarker for pregnancy outcomes. Thus, the observed suppression of *PAEP* gene expression and glycodelin secretion induced by 5 µM SE suggests a potential disruption of these critical immunomodulatory pathways.

As previously demonstrated, hormonally stimulated OFEL enhanced blastoid attachment, accompanied by increased secretion of total, β-, and hCG-h^40^. Our findings indicate that secretion of intact hCG and hCG-h was primarily driven by hormonal priming, whereas SE exerted selective, concentration-dependent effects on individual hCG isoforms. Specifically, 0.039 µM SE increased intact hCG secretion relative to the corresponding hormonally untreated control, consistent with an endocrine profile characteristic of differentiated syncytiotrophoblasts. At the highest concentration (5 µM), SE selectively increased free hCG-β secretion without affecting hCG-α or intact hCG. Given that hCG-α and hCG-β are encoded by the *CGA* and *CGB* gene loci, respectively, these findings suggest differential regulation of hCG subunit expression in response to GLP-1R activation. The increased hCG-h secretion observed under hormonally stimulated conditions is consistent with a hyperglycosylated hCG-dominant state characteristic of early implantation and invasive trophoblast differentiation^41^. Together with the observed effects on attachment assay, glycodelin secretion, and hCG secretion, these findings indicate that SE may modulate adhesive, immunomodulatory, receptivity-related, and hormonal aspects of endometrial biology, while still supporting implantation even in the absence of steroid hormone priming. However, as these findings are based on an in vitro blastoid attachment model, caution is warranted in extrapolating the results to in vivo settings, and definitive implications for reproductive outcomes in individuals using SE remain to be established.

To explore potential mechanisms of action, we next examined whether blastoids express GLP-1R, the canonical receptor mediating SE activity, and detected both protein and mRNA, with transcript levels remaining unchanged following SE treatment. While previous studies reported GLP-1R expression in mouse embryonic stem cells and post-implantation embryos^42^, its presence during pre-implantation stages has not been described. Single-cell RNA-seq resolved lineage-specific responses, demonstrating that ELC and TLC proportions were preserved following SE treatment. Given that the epiblast gives rise to all embryonic lineages and underpins pluripotency and inner cell mass integrity, whereas the trophectoderm contributes to most of the placenta and establishes fetal-maternal nutrient exchange, the observations following the SE treatment suggest that SE does not affect the cell type distribution in the pre-implantation blastoid model^43,44^. Within the epiblast and trophectoderm lineage, we observed coordinated upregulation of genes involved in glycolysis, oxidative phosphorylation, and the NRF2 antioxidant pathway, alongside downregulation of histone-modifying enzymes, consistent with metabolic remodeling and shift towards oxidative metabolism. Notably, oxidative phosphorylation activity increases during pre-implantation stages, rising from the day 5–6 blastocyst stage to day 6–7, before shifting dramatically upon implantation^45^. The observed downregulation of histone-modifying enzymes may reflect metabolic regulation of the epigenetic landscape, as previously suggested^46^. These findings suggest that GLP-1R activation drives adaptive metabolic and transcriptional changes in the human blastoids, paralleling physiological shifts in energy metabolism that occur during blastocyst maturation^45^, while also potentially intersecting with epigenetic regulation of the cell fate^46^.

In conclusion, our findings indicate that lower concentrations of SE have limited effects on the endometrial epithelium and stromal compartment, whereas higher concentrations enhance epithelial oxidative metabolism and exert predominant anti-proliferative effects on stromal cells. Across all concentrations, SE consistently impacts blastoids, highlighting potential consequences for embryo implantation and early development. This compartment-specific response creates a functional mismatch: the embryonic and epithelial compartments exhibit heightened metabolic activity, whereas the endometrial stroma becomes less supportive. Further evaluation of whole endometrial tissue from women undergoing GLP-1RA treatment could provide a broader view on the SE’s effects on endometrium. Given the critical role of stromal cells in establishing the immunomodulatory, tolerogenic environment required during early pregnancy, these observations underscore the need for further studies examining how SE may influence maternal immune components and early embryo–maternal crosstalk required for successful conception.

### Limitations of the study

A key consideration of the study is that 0.039 µM SE falls within the reported steady-state plasma range^47,48^, whereas 1.25 and 5 µM exceed clinically observed systemic concentrations. These higher concentrations were included to define concentration-dependent responses and are consistent with those commonly used in mechanistic in vitro studies of GLP-1 receptor agonists^49–51^. In addition, the receptor dependence of the stromal response remains unresolved, particularly given the lack of detectable GLP-1R expression in ESCs. Moreover, although the blastoid–endometrium co-culture captures key features of early embryo attachment, it does not recapitulate the full cellular, endocrine and immune complexity of implantation in vivo. Our models were derived from healthy, fertile donors and may not fully reflect responses in women with obesity, PMOS or metabolic dysfunction, who constitute partially the clinical population exposed to SE. Finally, the inferred metabolic reprogramming and changes in histone-modifying enzyme expression are based primarily on transcriptomic analyses and require direct functional validation.

## Methods

### Ethical approval

Endometrial biopsies of healthy fertile women in South Estonian Hospital (Võru, Estonia) were collected under ethical permissions No. 330/M-8 (16.10.2020) and No. 364/M-9 (16.05.2022) issued by the Research Ethics Committee of the University of Tartu. Ethical permits (454/02 and 2011/745/31-3) issued by Stockholm Region Ethical Review Authority allowed the use of embryonic stem cells to investigate human reproduction and blastoid generation.

### Participants and sample collection

Endometrial biopsies were obtained from healthy and fertile women (N=42) of reproductive age (32.4 ± 6.1 years old; body mass index 23.7 ± 3.5) using a Pipelle flexible suction catheter (Laboratoire CCD, Paris, France) (**Extended Data Table 5**). Inclusion criteria required participants to have at least one live-born child and to have abstained from hormonal medications for a minimum of three months prior to sample collection. Exclusion criteria included a history of sexually transmitted infections, uterine pathologies, endometriosis or PMOS. Menstrual cycle phase was determined based on self-reported menstrual cycle history. The proliferative phase (P) was identified based on menstrual cycle history. Early secretory (ES), mid-secretory (MS), and late secretory (LS) cycle phases were determined by counting days after the luteinizing hormone (LH) surge, detected using the BabyTime hLH urine cassette (Pharmanova, Belgrade, Serbia). Specifically, days LH+2 and LH+3 were classified as ES, LH+7 to LH+9 as MS, and LH+11 to LH+12 as LS.

Immediately following collection, the biopsies were transferred into HypoThermosol FRS Preservation Solution (Sigma, St. Louis, MO, USA). Each biopsy was bisected: one portion was fixed in 10% formalin for histological evaluation, while the other was preserved in a cryopreservation medium comprising 1× Dulbecco’s Modified Eagle’s Medium (DMEM; Thermo Fisher Scientific), 30% fetal bovine serum (FBS; Biowest, Riverside, MO, USA), and 7.5% Hybri-Max Dimethyl Sulfoxide (Sigma-Aldrich). Cryopreserved samples were placed in the Nalgene Cryo 1°C ‘Mr. Frosty’ freezing container (Thermo Scientific) and stored at −80 °C overnight before being transferred to liquid nitrogen for long-term storage.

### Establishing paired endometrial epithelial organoids and stromal cell culture

Cryopreserved endometrial tissue samples were thawed rapidly in a 37 °C water bath. Once fully thawed, tissues were immediately transferred to 15 ml conical tubes and washed twice with 5–7 ml of pre-warmed DMEM (phenol red–free; Corning), supplemented with 10% charcoal-stripped FBS and antibiotic–antimycotic solution to remove residual cryoprotectant and blood contaminants. Tissue dissociation was conducted by enzymatic cocktail composed of 1.25 U/ml Dispase II (Sigma-Aldrich, D4693) and 0.4 mg/ml Collagenase V (Sigma-Aldrich, C9263). The samples were first agitated manually, then incubated at 37 °C in a horizontal position on a rotating shaker for up to 20 min, with vigorous shaking every 5 min to enhance dissociation. Following enzymatic digestion, the cell suspension was passed through a 100-μm cell strainer to remove undigested tissue and debris. The flow-through, enriched for endometrial stromal cells, was centrifuged and the resulting pellet resuspended and cultured in phenol red–free DMEM (Gibco) supplemented with 10% charcoal-stripped FBS (Gibco) and penicillin/streptomycin/amphotericin B (100 U/ml, 100 μg/ml, and 0.25 μg/ml, respectively), at 37 °C and 5% CO₂. The retained fraction from the cell strainer, containing endometrial glands, was used to generate endometrial epithelial organoids following established protocols (**Fig. 1a**)^52,53^.

Before the exposures the EEOs were dissociated into single cells and seeded at 10,000 cells per well. Briefly, EEO in 48-well plate were recovered from Matrigel (Corning) by treating with 25 μL of Dispase (10 mg/mL) for 1 hour at 37 °C, then transferred to a 1.5 mL low-binding tube and centrifuged at 600g for 5 min. After washing with Advanced DMEM/F12 + Y-27632 (10 nM), they were resuspended in 1 mL dissociation solution (TrypLE, N-acetylcysteine [51 μg/mL], and Y-27632 [10 nM]) and incubated at 37 °C for 15 min, with periodic pipetting to monitor dissociation. Cells were counted (Corning Cell Counter) and seeded in 10,000 cells per well in 48 well plate.

### Hormonal stimulation and semaglutide treatment

To recapitulate the molecular profile of endometrial receptivity, endometrial epithelial cells were treated with 10 nM β-estradiol (E2; Sigma) for 48 hours. Cells were then further treated with either (i) 10 nM E2 alone or (ii) a combination of 10 nM E2, 1 μM progesterone (P4; Sigma), and 1 μM 8-bromoadenosine 3′,5′-cyclic monophosphate (8-Br-cAMP; Sigma) for an additional 4 days, as previously described^54^. For in vitro decidualization of ESCs, the protocol was carried out as described^55^. ESCs were exposed to 10 nM E2, 100 nM P4, and 200 μM 8-Br-cAMP for 9 days, with media, with or without hormones, replaced every 72 hours. SE (Ozempic, Novo Nordisk) stock was prepared at 328 μM, and treatments consisted of 0.039 μM, 1.25 μM, and 5 μM SE, either alone or in combination with the hormonal cocktail.

### Endometrial stromal cells decidualization markers

The secretion of PRL and IGFBP-1 was measured in culture media samples collected during the final 72 hours of hormonal treatment. PRL was quantified using a PRL ELISA kit (Cayman Chemical, Ann Arbor, MI, USA), and IGFBP-1 was quantified using an IGFBP-1 ELISA kit (Invitrogen, Frederick, MD, USA), following the manufacturers’ protocols. IGFBP-1 samples were diluted 2.1- to 41-fold depending on the sample. For each tested sample, the concentration of PRL was determined from a calibration curve using linear regression, whereas IGFBP-1 concentrations were determined from a calibration curve using non-linear regression with a one-phase association equation. For samples obtained from the same donor, data were normalized to the E2 treatment (0% secretion) and the EPC treatment (100% secretion). Normalized data were then pooled for each treatment group (PRL: 3 women, samples in duplicates; IGFBP-1: 4 women, samples in duplicates).

### Relative metabolic activity assay

To assess the metabolic activity of EEO and ESC, we performed a resazurin assay as previously described^56^. Briefly, EEO were dissociated into single cells, and 4,000 cells per well were seeded onto mini ring-shaped Matrigel in 96-well Flat Clear Bottom White Polystyrene TC-treated microplates (Corning). The mini-ring shape facilitates high-throughput compound testing^57^. After Matrigel solidification, 100 µL of media were added to each well. The cell culture was maintained until organoids formed, typically within 4 to 5 days. Once organoid formation was complete, exposure treatments were initiated. For ESCs, 5,000 cells were seeded onto 96-well Flat Clear Bottom White Polystyrene TC-treated microplates (Corning). The exposure treatments consisted of SE at concentrations of 0.039 µM, 1.25 µM, and 5 µM for 6 days in EEOs and 9 days in ESCs. The incubation times were selected to assess the effects on metabolic activity in response to hormonal stimulation, with a 6-day treatment for EEO and a 9-day treatment for ESC.

After incubation, the growth medium was removed, the cells were rinsed with DPBS (Capricorn), and 50 µM resazurin (Sigma) solution in DPBS supplemented with Ca2+ and Mg2+ (Capricorn) was subsequently added. The plates were placed into multi-mode reader Cytation 5 (BioTek), and measurement of absorbance was performed (570 nm and 600 nm, monochromator; kinetic mode with reading taken every 15 min for 2 hours, read height 8.5 mm, with lid).

For data analysis, ratio of absorbance at 570 nm and 600 nm was calculated for each well and time-point. Next, the fluorescence intensity-derived data and the absorbance ratio-derived data were analyzed separately and considered as biological replicates. For each individual patient’s cells, the data obtained for the negative control incubations (non-treated cells) was pooled and plotted against time, and the linear range of the assay was established. For the chosen time-point, the data for treated cells was normalized according to the negative control (100% metabolic activity) and positive control (no cells seeded, 0% metabolic activity). The normalized data was then pooled for the same kind of treatment (3 patients, samples in sextuplicates for each treatment).

### Seahorse oxygen consumption measurement in EEO

OCR was measured using the XF24e extracellular flux analyzer (Seahorse, Agilent) with the Mito Stress Test. Briefly, four wells of EEOs were washed with ice-cold 1× PBS (Corning) and centrifuged at 500 × g for 6 min to remove Matrigel. Subsequently, 3 μL of Matrigel containing EEOs was seeded into XF24-well plates (Agilent) in 200 μL of expansion medium. Organoids were cultured for a minimum of 2 days prior to the Seahorse assay to allow the recovery of morphology. EEOs were treated with SE (0.039 µM, 1.25 µM and 5 μM) following short (40-minute) and prolonged (6-day) protocol. Before the assay, organoids were washed twice with 500 μL of Seahorse XF DMEM medium supplemented with 10 mM glucose, 5 mM pyruvate, and 2 mM glutamine, and incubated at 37 °C in a non-CO₂ incubator for 45 min. Compounds were loaded into the cartridge in the following order and final concentrations: port A - SE (0.039 µM, 1.25 µM and 5 μM), port B - oligomycin (1 μM), port C - FCCP (1 μM), and port D - rotenone (1 μM) and antimycin A (1 μM). Following cartridge calibration, the cell plate was loaded and OCR measurements were performed. Each experiment included three biological replicates and five technical replicates. Final data were normalized to DNA content measured using a NanoDrop spectrophotometer.

### cAMP detection

Activation of GLP1R in response to SE stimulation in EEO and ESCs was assessed by measuring intracellular cAMP levels, which serve as a second messenger regulating downstream signaling pathways. EEOs were dissociated into single cells as previously described, and 35,000 epithelial cells were seeded per well. ESCs were seeded at a density of 5,000 cells per well. 96-well Flat Clear Bottom White Polystyrene TC-treated microplates (Corning) were used for the assay. Two days after seeding, EEOs were treated with 10 nM E2 and 1 μM P4 beginning 2 hours after seeding or left unstimulated and maintained for 6 days with media changes every 2 days. ESCs were treated with 10 nM E2 and 100 nM P4 for 9 days or left unstimulated, with media changes every 2 days.

The concentration of intracellular cAMP was measured using the BacMam compatible version of the biosensor expression vector^58^ mTurq2Δ_Epac(CD,ΔDEP, Q270E) td ^cp173^Ven (H187) according to the previously reported protocol^59^, with minor modifications. Briefly, viral stock was prepared in fresh growth medium containing 10 mM sodium butyrate (Sigma-Aldrich) for enhanced protein expression and the solution was added onto the cells (100 mL per well, final multiplicity of infection: 100). The cells were grown for 48 h at 37 °C in a humidified CO2 incubator. On the day of the assay, the growth medium was replaced with 100 μL phosphate-buffered saline (PBS) one hour prior to the experiment. The dilutions of SE or positive control forskolin (Tocris Bioscience) on transparent 96-well clear-bottom plates (Thermo Fisher Scientific) were made into PBS containing 7.5 mol/L BSA (Sigma-Aldrich) to reduce non-specific binding. The fluorescence intensities were registered prior and after addition of the compounds using PHERAstar plate reader (BMG LABTECH GmbH), with excitation at 427 nm and simultaneous dual emission at 480 (CFP channel) and 530 nm (YFP channel). The final concentrations of compounds on the cells were 30 µM in case of forskolin and 0.039 µM, 1.25 µM or 5 µM in case of SE. After addition of compounds, readings were taken every 30 seconds for 60 min. The change in FRET ratio (ΔFRET) for each time-point was calculated by subtracting the ratio of YFP and CFP channels at the given time-point from the corresponding ratio at the time zero (prior to addition of the compounds) and dividing the obtained difference with the channel ratio at the time zero. For the chosen time-point (10 min or 60 min after addition of compounds), the data for treated cells was normalized according to the positive control (100% DFRET) measured in the same patient’s cells. The normalized data was then pooled for the same kind of treatment (3 women, samples in pentaplicates for each treatment).

### Blastoids generation

Naive human embryonic stem cells (hESCs) H9 were cultured on gelatin-coated plates including a feeder layer of gamma-irradiated mouse embryonic fibroblasts (MEFs) in PXGL medium. PXGL medium is prepared using N2B27 basal medium supplemented with PD0325901 (1 µM, MedChemExpress, HY-10254), XAV-939 (2 µM, MedChemExpress, HY-15147), Gö 6983 (2 µM, MedChemExpress, HY-13689) and human leukemia inhibitory factor (hLIF, 10 ng/ml, Proteintech Group). N2B27 basal medium contained DMEM/F12 (50%, Gibco/Life technologies), neurobasal medium (50%, Gibco/Life technologies, 21103049), N-2 supplement (Thermo Fisher Science, 17502048), B-27 supplement (Thermo Fisher Science, 17504044), GlutaMAX supplement (Thermo Fisher Science, 35050-038), and 2-mercaptoethanol (100 µM, Thermo Fisher Science, 31350010). Cells were routinely cultured in hypoxic incubator (5% CO2, 5% O2). Cell lines had routinely been tested negative for mycoplasma.

For generating blastoids, naïve H9 hESCs were cultured and passaged 4 days before blastoids formation. Blastoids were initiated as previously described^60^. In brief, naïve H9 cells were treated with Accutase at 37 °C for 5 min, followed by gentle mechanical dissociation by pipette. The dissociated cells were centrifuged and pelleted cells were resuspended in PXGL medium with Y-27632 (10 μM, MedChemExpress, HY-10583). The cell suspension was plated onto 0.1% gelatin-coated plate and was incubated for 1 hour at 37 °C to exclude MEFs. After MEFs exclusion, the cell number was estimated by using automated cell counter. Then cells were resuspended in aggregation medium (N2B27 medium containing 10 mM Y-27632 and 0.4% BSA) to seed 100,000 cells per well in AggreWell 400 24-well plate (StemCell Technology). The plate was placed in a hypoxic incubator (5% CO2, 5% O2) for the whole period of blastoid formation. The cells were allowed to form aggregates inside the microwell for 24 h (day 1). Subsequently on day 2, the aggregation medium was replaced with 2x PALLY medium (N2B27 supplemented with PD0325901 (1 µM), A 83-01 (1 µM, MedChemExpress, HY-10432), 1-oleoyl lysophosphatidic acid sodium salt (LPA) (2 µM, Tocris, 3854), hLIF (10 ng/ml), 0.3% BSA (Sigma) and Y-27632 (10 µM)). The PALLY medium was refreshed on day 3. On day 4, the PALLY medium was replaced with N2B27 medium containing 2 µM LPA, 0.4% BSA, and 10 µM Y-27632. On day 5, the blastoid formation rate was estimated based on cavity formation and expansion. The blastoids were collected on day 6, i.e., 120 hours.

For blastoids, SE was added to the culture media on day 4 (0.039 µM and 5 µM). On day 5, the media was refreshed, and the wells were imaged. On day 6, the blastoid formation efficiency was assessed based on morphology, and images were acquired.

### *In vitro* attachment assay

To evaluate the potential effect of SE on blastoids attachment to endometrial epithelial cells, the in vitro attachment assay was performed as previously described^60,61^. The EEOs were dissociated into single cells as previously described and seeded at a confluency of 35,000 cells per well into 3%-Matrigel pre-coated 96-well clear bottom plate (Corning) and immediately stimulated with E2 (10nM) for two days, followed by the mixture of E2 (10 nM), P4 (1 μM), cAMP (250 μM) and XAV939 (10 μM) for 4 days (**Fig. 1b**). For blastoids transfer, CMRL1 media containing embryonic stem-cell FBS (10%), GlutaMAX (2mM), N2 (1x), B27 (1x), sodium pyruvate (1mM), E2 (10nM), P4 (1 µM) and Y-27632 (10 µM) were added to the OFEL 2 hours before blastoid transfer. Five blastoids per well were added with a stripper (CooperSurgical, USA) (**Fig. 1b**). After 48 h, plates were washed with PBS, and the evaluation of the attachment was quantified as the percentage of attached blastoids versus the five blastoids initially transferred (**Fig. 1b**). Media was collected for further hCG and glycodelin detection.

### hCG and glycodelin immunoassays

Immunofluorometric assays were used to quantify the secretion of hCG and glycodelin in the media from in vitro blastoid attachment assays. The different forms of hCG measured in the culture media included intact hCG (αβ heterodimer, lowest standard 2.12 IU/L), free α subunit (hCGα, lowest standard 1.5 pmol/L), free β subunit (hCG-β, lowest standard 0.34 pmol/L), and hyperglycosylated intact hCG (hCG-h, lowest standard 8.75). Glycodelin concentration (lowest standard 1.08 ng/mL) was also assessed to evaluate endometrial receptivity. Prior to analysis, media were diluted 1:50 for hCG-isoform and subunit assays, and 1:3 for glycodelin. Intact hCG levels were detected using a commercial immunoassay (Perkin Elmer Wallac, Finland), while free hCG-α, free hCG-β, hCG-h, and glycodelin were measured using established in-house immunoassays: free hCG-α^62^, free hCG-β^63,64^, hCG-h^65^, and glycodelin^66^.

### RNA extraction, library preparation, sequencing and data analysis

Gene expression changes induced by SE treatment in EEO and ESCs were assessed by bulk RNA sequencing. Endometrial epithelial cells derived from dissociated EEO were seeded at 10,000 cells per well in 48-well plates. Upon organoid formation, hormonal and SE treatments were applied for 6 days, as previously described. ESCs were seeded at 20,000 cells per well in 48-well plates and treated with hormones and SE for 9 days following the in vitro decidualization protocol.

RNA was extracted using the RNeasy Micro Kit (Qiagen) with DNase treatment, and RNA concentration was quantified using a NanoDrop spectrophotometer (Thermo Fisher, USA). RNA samples were stored at –80 °C until further processing. Libraries were prepared using the TruSeq Stranded mRNA kit (Illumina, USA) and sequenced on the NextSeq 1000 platform (Illumina, USA) with single-end 80 bp setting.

Raw sequencing reads were pre-processed using FastQC and Trim Galore and aligned to the human genome version 38 (GRCh38) using HISAT2 within the Galaxy platform^67^. Differential expression analysis was performed with DESeq2. DEGs were identified using thresholds of |log₂FC| ≥ 1 for EEOs and ESCs, with adjusted p-value (Padj) < 0.05. Heatmaps were generated using Morpheus (https://software.broadinstitute.org/morpheus), and gene ontology (GO) enrichment analysis was conducted using ShinyGO^68^. To assess whether SE produced a concordant transcriptional response across decidualization contexts, gene-level log₂ fold changes (SE vs. control) from the DESeq2 analyses were compared between conditions for all commonly detected genes, and their agreement was quantified using Pearson’s correlation coefficient (r).

### scRNA-seq

### Single-cell dissociation of blastoids

Human blastoids were cultured and subjected to treatment beginning on day 4 of development. Three experimental conditions were established: untreated control, and treatment with 0.039 μM, and 5 μM SE. SE (Semaglutide, Ozempic®, Novo Nordisk, Denmark) was added to the culture media on day 4 (**Fig. 1c**). On day 5, an equal volume of fresh medium was added to each well without replacing the existing medium.

For scRNA-seq, blastoids were collected on day 6. A total of 20 wells per condition were harvested from the AggreWell™ plate (STEMCELL Technologies, Canada) and pooled into 15 mL conical tubes (10 wells/tube, 2 tubes per condition). Due to the large volume and size of the blastoids, passive sedimentation was not effective; instead, samples were centrifuged at 200 x g for 2 min and the supernatant was discarded. To initiate single-cell dissociation, 3 mL of Accutase (Innovative Cell Technologies, USA) was added to each pooled sample (10 wells), followed by gentle pipetting (10 times) using a 1 mL pipette. Samples were incubated at 37 °C for 10 min, with mechanical dissociation performed every 5 min by pipetting (10–15 times) under microscopic observation to ensure optimal single-cell dissociation. Following enzymatic digestion, cells were centrifuged at 300 × g for 5 min and washed once with PBS containing 0.04% BSA to remove ambient RNA. The cells were resuspended in 0.04% BSA in DPBS to attain a concentration of 700-1,200 cells/μl as per the 10x Genomics protocol.

### Chromium 10x single-cell capturing, library generation and sequencing

Single-cell suspensions derived from treated and untreated blastoids were processed for single-cell RNA sequencing using the 10x Genomics Chromium platform. The single-cell suspension for all three conditions (targeting 5,000 cells/condition), single-cell 3′ gel beads (3’ Gel Bead Kit v4 4-plex), and reverse transcription master mix were loaded onto a 10x Chromium microfluidic 3’ OCM Chip (GEM-X OCM 3’ Chip Kit v4 4-plex). A multiplexed, pooled Gel Beads-in-Emulsion (GEMs) (20,000 cells in total) was generated using the Chromium Controller X.

Library construction was performed using the Library Construction Kit C and Dual Index Kit TT Set A (10X Genomics, USA), following the manufacturer’s protocol. The workflow included cell lysis, reverse transcription, barcoding, cDNA amplification, and purification to generate full-length barcoded cDNA from individual cells. Quality control of amplified cDNA and final dual-indexed libraries was performed using the Agilent 4150 TapeStation (Agilent Technologies, USA). Libraries were then pooled and sequenced on a NovaSeq X Plus Series platform with paired-end 150 bp reads (PE150), targeting a sequencing depth of approximately 30,000 reads per cell.

### Pre-processing single-cell RNA-seq data

Raw scRNA-seq FASTQ files from control blastoids and blastoids treated with 0.039 μM and 5 μM of SE were aligned to the human reference genome GRCh38 (v3.0.0, downloaded from the 10X Genomics website) using the CellRanger (v9.0.1) with default settings for the 10X Genomics pipeline. Low-quality cells were filtered based on the number of expressed genes (nGene) and the percentage of mitochondrial gene expression (percent.mito). Only cells with 750 < nGene < 5,000 and percent.mito < 0.2 were retained for downstream analysis. Following quality control and exclusion of mitochondrial genes, we retained genes with at least one count in five or more cells.

Log-normalized expression values were computed using the deconvolution strategy implemented in the computeSumFactors function from the scran R package (v1.30.2)^69^. These values were further normalized across treatment groups using the multiBatchNorm function from the batchelor R package (v1.18.1)^70^ to ensure comparability of size factors. The rescaled, log-normalized expression values were subsequently used for dataset dimensionality reduction and differential gene expression analysis.

### Dimensionality reduction and clustering

To perform dimensionality reduction and clustering, we processed the single-cell gene expression matrix using the Seurat package (v5.1.0)^71^. Variable genes were identified using the FindVariableFeatures function with the “vst” method, selecting the top 2,000 most variable features. The data were then scaled using the ScaleData function, regressing out the percentage of ribosomal gene expression to reduce technical noise. Principal component analysis (PCA) was performed using the RunPCA function, and the top 20 principal components were used for further analyses. Uniform manifold approximation and projection (UMAP) was applied using the RunUMAP function^72^ to visualize the data in two dimensions. Finally, a nearest-neighbor graph was constructed using the FindNeighbors function based on the first 20 principal components to facilitate downstream clustering and analysis. The predicted lineage scores for each cell were calculated using the AddModuleScore function from Seurat package, incorporating the top 15 identified lineage marker genes^18^.

### Detection of marker genes and differentially expressed genes from scRNA-seq data

Marker gene detection was performed using the FindAllMarkers function in the Seurat package. Differential gene expression analysis was conducted using the Wilcoxon rank-sum test, implemented in Seurat’s FindMarkers function. Genes were considered significantly differentially expressed if they had a false discovery rate (FDR) < 0.05, a log2 fold change > 0.25, and were expressed in more than 15% of cells. Genes that were consistently upregulated or downregulated across all treatment groups and were significantly differentially expressed in at least one treatment condition were defined as consistently up- or down-regulated DEGs.

### Projection onto human early embryo reference using scRNA-seq data

Projection onto a human early embryogenesis reference dataset was performed using the Early Embryogenesis Projection Tool (v2.1.2)^18^ (available at: https://petropoulos-lanner-labs.clintec.ki.se). Predicted UMAP coordinates and cell-type annotations were downloaded from the tool for visualization.

### Metabolic activity prediction from scRNA-seq data

ScRNA-seq data from the SE treated blastoids and controls were analyzed using the previously published scCellFie pipeline ^19^ to infer single-cell metabolic activity.

### Immunoffuorescence staining

Human blastoids were fixed with 4% paraformaldehyde (PFA) in PBS for 30 min at room temperature and washed with 1X PBS. The fixed blastoids were permeabilized with 0.5% Triton X-100 in PBS for 30 min at room temperature. Blastoids were blocked with blocking solution (PBS containing 5% BSA, 0.1% Tween-20 and 0.01% TritonX-100) at room temperature for 1 h. Then, blastoids were incubated with primary antibodies for OCT3/4 (sc-5279, Santa Cruz Biotechnology), GATA3 (sc-9009, Santa Cruz Biotechnology), and GATA4 (14-9980-82, eBioscience, Invitrogen) diluted (1:200) in washing buffer and incubated overnight at 4 °C. Blastoids were washed three times with washing buffer under gentle orbital shaking for 5 min each, followed by incubation with Alexa 488, Alexa 555 and Alexa 647, diluted (1:1,000) and Hoechst (1:1,000) in washing buffer for 1h at room temperature. Blastoids were washed three times with washing buffer as described above. Confocal imaging was performed in 16 wells m-slides (Ibidi) on a Nikon Ti2 spinning-disk confocal microscope (**Fig. 1c**).

The EEOs were incubated with Cell Recovery Solution (Corning) for 30 min and washed with cold Advanced DMEM (Thermo Fisher) to remove Matrigel. They were then fixed with 4% PFA for 45 min and washed three times with PBT (0.01% Tween-20 in 1× PBS). Blocking was performed using a blocking solution (10% donkey serum, 0.1% Triton X-100, and 0.2% BSA in 1× PBS) for 15 min at 4 °C. The EEOs were incubated overnight at 4 °C with primary antibodies: GLP-1R (MAB7F38, DSHB 1:30) and Ki-67 (sc-23900, Santa Cruz, 1:100), diluted in washing buffer (0.1% Triton X-100 and 0.2% BSA in 1× PBS). After three washes, they were incubated with secondary antibodies, Alexa Fluor 488 (1:1,000) and Alexa Fluor 647 (1:1,000) for 1 hour at room temperature. The EEOs were washed three more times before imaging with a Nikon Ti2 spinning-disk confocal microscope.

### Immunohistological evaluation of human endometrial tissue samples

For immunohistochemical analysis, 2.5 µm sections were prepared at the Pathology Department of Tartu University Hospital (Tartu, Estonia) from formalin-fixed, paraffin-embedded (FFPE) endometrial tissue. Sections were deparaffinised and rehydrated through graded ethanol into water, followed by heat-mediated antigen retrieval at 98 °C for 20 min in 10 mM sodium citrate buffer (pH 6.0). For permeabilisation, sections were incubated in 0.1% Triton X-100 (in PBS) for 20 min, blocked for 1 h in 5% normal goat serum, and then incubated overnight at 4 °C with primary antibody: GLP-1R (MAB7F38, DSHB) diluted 1:30 in 1% BSA/PBS. Negative controls were processed identically but without the primary antibody. Human pancreatic tissue was used as a positive control for GLP-1R immunostaining. After washing in PBS, sections were incubated with goat anti-rabbit Alexa Fluor 488 secondary antibody (1:1000; Thermo Fisher Scientific) for 1 h at room temperature, counterstained with DAPI (4′,6-diamidino-2-phenylindole; Sigma-Aldrich), washed in PBS, and mounted in Fluoromount-G (Thermo Fisher Scientific). Slides were stored at 4 °C until imaging. Images were acquired on an Olympus BX51 microscope with a 20× objective using Cell B acquisition software (Olympus).

Images were analysed in Fiji (ImageJ 1.54p). Quantification used the AF488 grayscale channel (no RGB composites). Glandular epithelium (Epi ROI) was obtained by outlining the gland and lumen and computing the epithelial ring with ROI Manager (XOR); ROIs were saved. Epithelial area (µm²) was measured with Limit to Threshold off. For GLP-1R–positive area, a numeric lower threshold (T) was set per imaging session on the matched negative control so that a small stromal background ROI yielded an area fraction ≤1%. With Limit to Threshold on, the GLP-1R–positive epithelial area fraction (%) was measured within the Epi ROI as: positive area (µm²) = Epi area × (area fraction/100). Statistics were performed in GraphPad Prism 8.0.1 (GraphPad Software). For each sample, gland-level measurements were summarised as the per-sample median GLP-1R–positive epithelial area fraction (%). Differences across menstrual phases were tested with the Kruskal–Wallis test. Pairwise comparisons used Dunn’s multiple comparisons with multiplicity-adjusted p values. Data are presented as per-patient values shown as violin plots with group medians and interquartile range (IQR). All tests were two-sided with α = 0.05, and groups were analysed as unpaired.

### Quantitative RT-PCR analysis of GLP-1R expression

mRNA expression levels of *GLP-1R* (assay ID: Hs00157705_m1) and the housekeeping gene *TBP* (assay ID: 4333769F) were quantified using TaqMan™ probe-based assays with TaqMan™ Fast Advanced Master Mix (Thermo Fisher Scientific), according to the manufacturer’s instructions. DNase-treated total RNA (up to 1 μg), extracted from endometrial tissues at P (n = 5), ES (n = 6), MS (n = 8) and LS (n = 3) phases, was reverse transcribed into cDNA using the RevertAid First Strand cDNA Synthesis Kit (Thermo Fisher Scientific, MA, USA). cDNA was also synthesized from RNA isolated from SE-treated ESCs and EEO cultures stimulated with E2, EPC and untreated controls. qRT-PCR was performed for all these reactions using a Bio-Rad qPCR system (CFX Opus 96). Gene expression was normalized to *TBP*, and relative *GLP1R* expression levels (ΔCT) were compared with respective controls using Student’s two-tailed t-test. A p-value < 0.05 was considered statistically significant. Pancreatic β-islet organoids served as a positive control to validate the assay.

## Data Availability

All data produced in the present study are available upon reasonable request to the authors.

## Extended Data Figures

**Extended Data Fig. 1.**
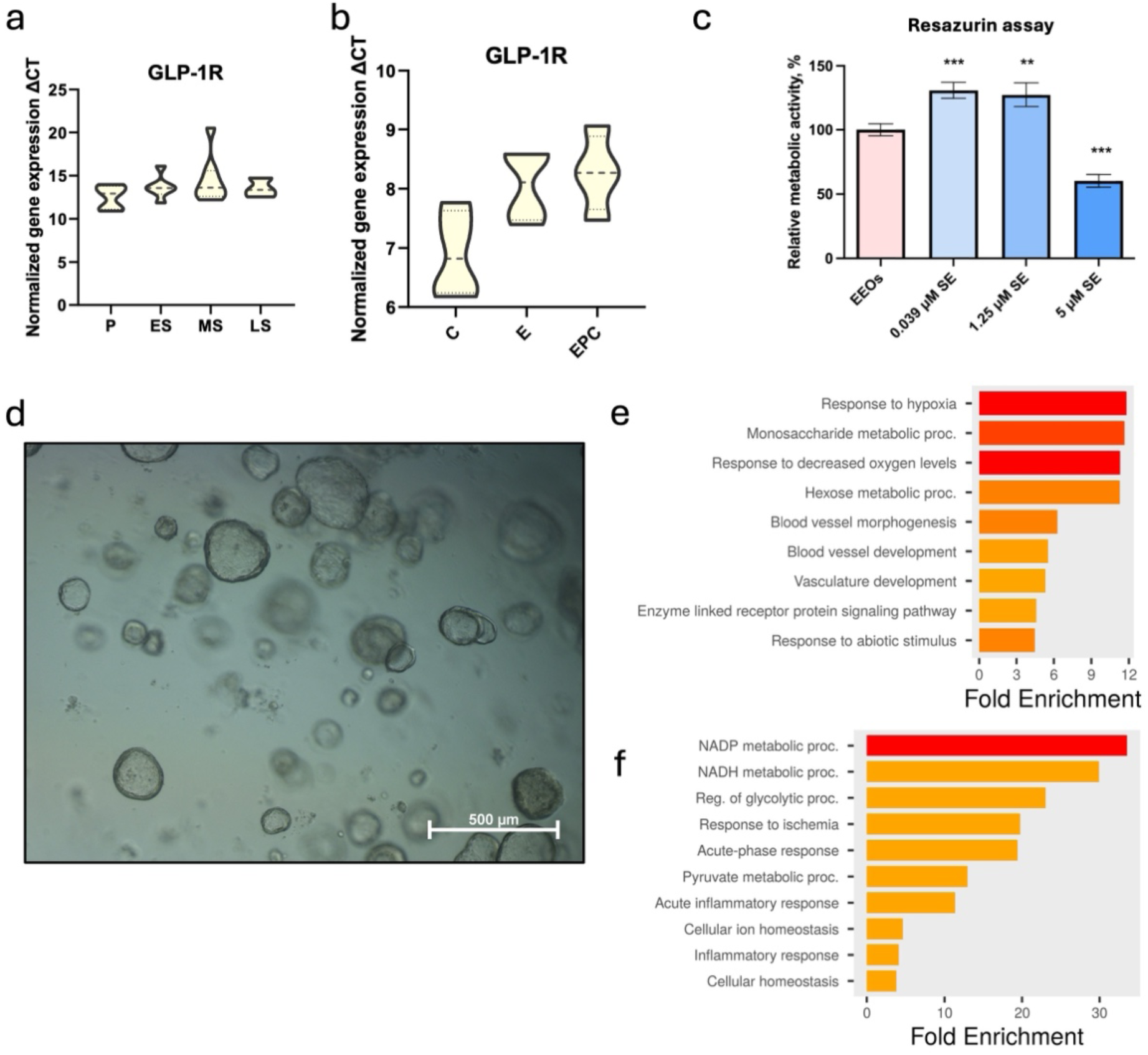
Expression and functional responses to semaglutide (SE) in human endometrial epithelial cells. **a**, *GLP1-R* gene expression measured by qPCR in endometrium from healthy fertile women, representing proliferative (P, N=5), early secretory (ES, N=6), mid-secretory (MS, N=8), and late secretory (LS, N=3) cycle phases. **b**, *GLP1-R* expression measured by qPCR in endometrial epithelial organoids (EEOs) from four biological replicates as untreated control (C), estrogen (E2), and estrogen + progesterone + cAMP (EPC) treatment conditions. **c**, Relative metabolic activity of SE-treated (0.039 μM, 1.25 μM, or 5 μM) EEOs and SE-untreated EEOs (control) assessed by resazurin assay. Error bars represent the Standard Error of the Mean (SEM) calculated from three independent biological replicates, with each experiment performed in technical triplicates. Statistical significance was determined using a one-way ANOVA followed by Dunnett’s multiple comparisons test (**p < 0.01, ***p < 0.001). **d**, Bright-field representative image of EEOs after 6 days of treatment with 5 μM SE. **e,** Gene ontology (GO) biological pathway enrichment of downregulated differentially expressed genes (DEGs) in EPC + SE 5 μM versus EPC alone in EEOs. **f**, GO biological pathways enrichment of upregulated DEGs in EPC + SE 5 μM versus EPC in EEOs.

**Extended Data Fig. 2.**
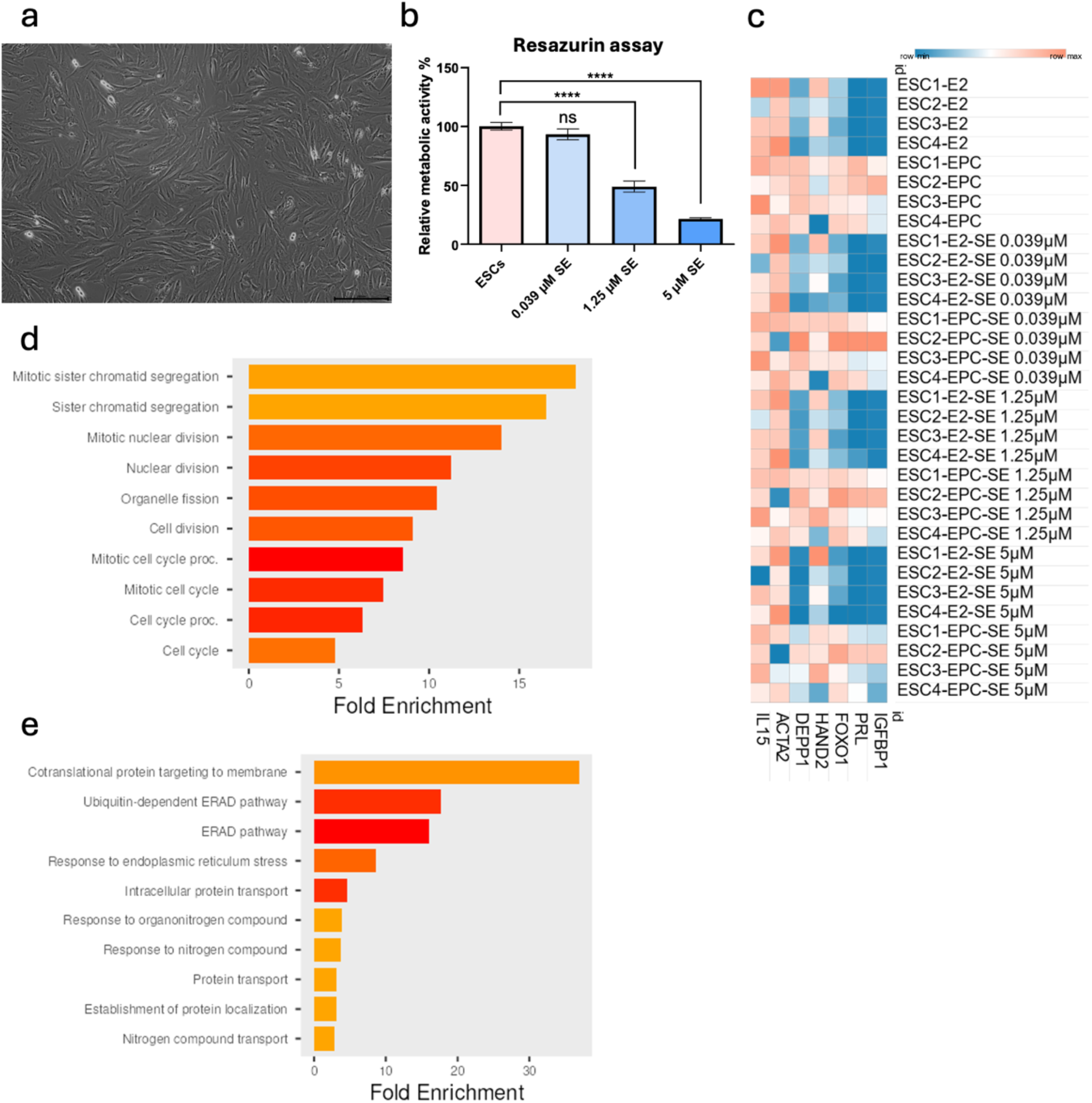
Expression and functional responses to semaglutide (SE) in endometrial stromal cells (ESCs). **a**, Bright-field representative image of endometrial stromal cells (ESCs) after 6 days of treatment with 5 μM SE. **b,** Relative cellular metabolic activity of SE-treated (0.039 μM, 1.25 μM, or 5 μM) and untreated ESCs assessed by resazurin assay. **c**, Heatmap of decidualization marker expression (log₂-transformed normalized counts) in ESCs across treatment conditions: estrogen (E2) alone, estrogen + progesterone + cAMP (EPC) without SE, E2 with SE (0.039 μM, 1.25 μM, or 5 μM), EPC with SE (0.039 μM, 1.25 μM, or 5 μM) (log₂-transformed normalized counts). **d**, Gene ontology (GO) biological pathways enrichment of downregulated differentially expressed genes (DEGs) in ESCs treated with EPC + SE 5 μM versus EPC alone. **e**, GO biological pathways enrichment of upregulated DEGs in ESCs treated with EPC + SE 5 μM versus EPC alone.

**Extended Data Fig. 3.**
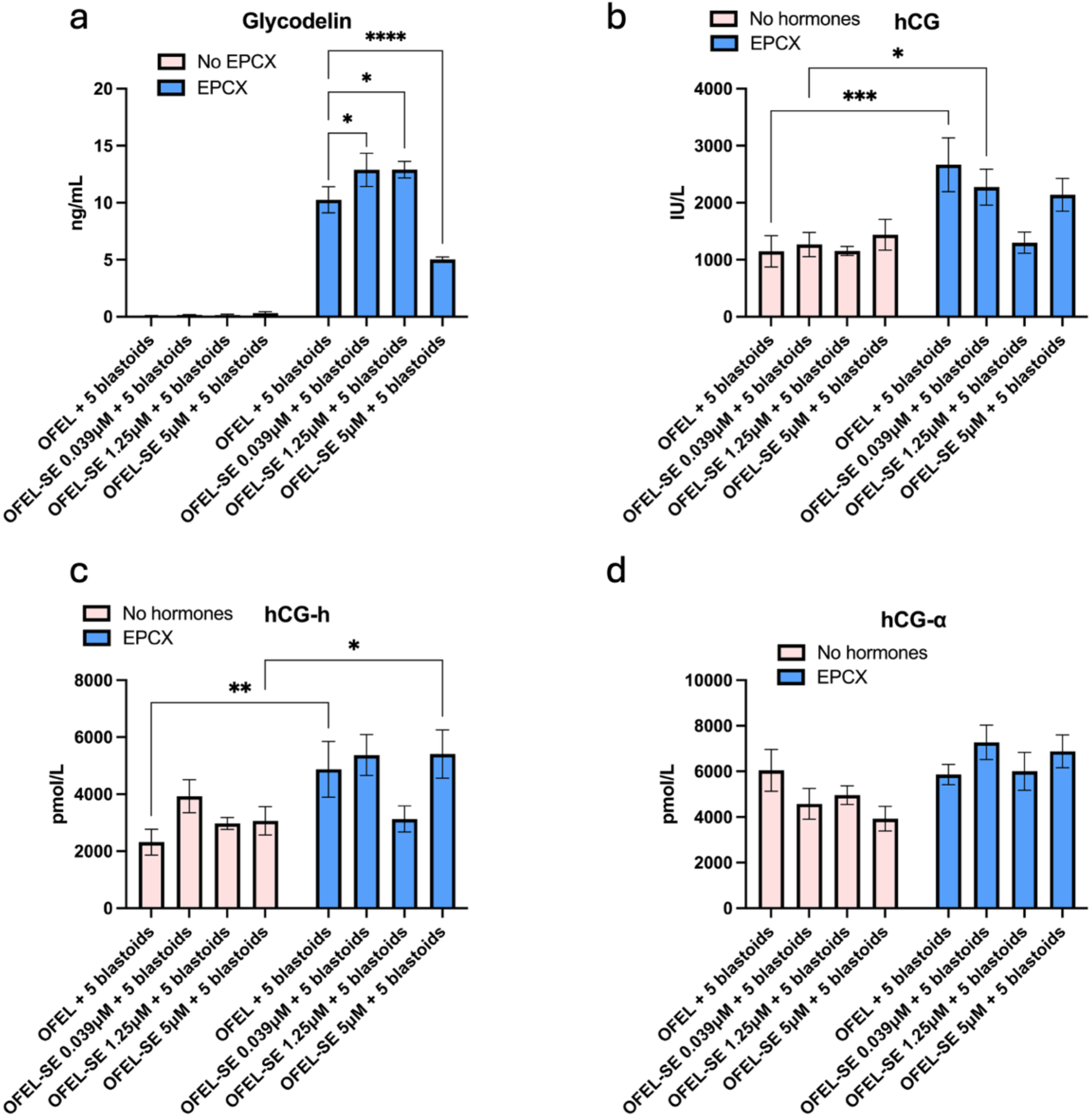
Glycodelin and human chorionic gonadotrophin (hCG) secretion in the blastoid attachment assay following semaglutide (SE) treatment. **a**, Glycodelin concentrations (ng/mL), measured by time-resolved fluorescence (TRF) immunoassay, in spent culture media from open-faced endometrial layer (OFELs) co-cultured with 5 blastoids per well and treated with SE (OFEL-SE-0.039 μM, 1.25 μM, or 5 μM), in the absence and presence of EPCX (estrogen, progesterone, cAMP, and XAV-939) conditions. **b**, Total hCG levels (IU/L), measured by TRF immunoassay in the same blastoid attachment conditions as described in A. **c**, hyperglycosylated hCG and **d**, hCG-α subunit levels (pmol/L), both measured by TRF immunoassay in spent media from the same experimental conditions. Error bars represent the SEM calculated from two biological replicates, with two independent experiments (N=4). Each experiment was performed in five technical replicates. Statistical significance was determined using a two-way ANOVA followed by Tukey’s multiple comparisons test (***p < 0.001, ****p < 0.0001).

**Extended Data Fig. 4.**
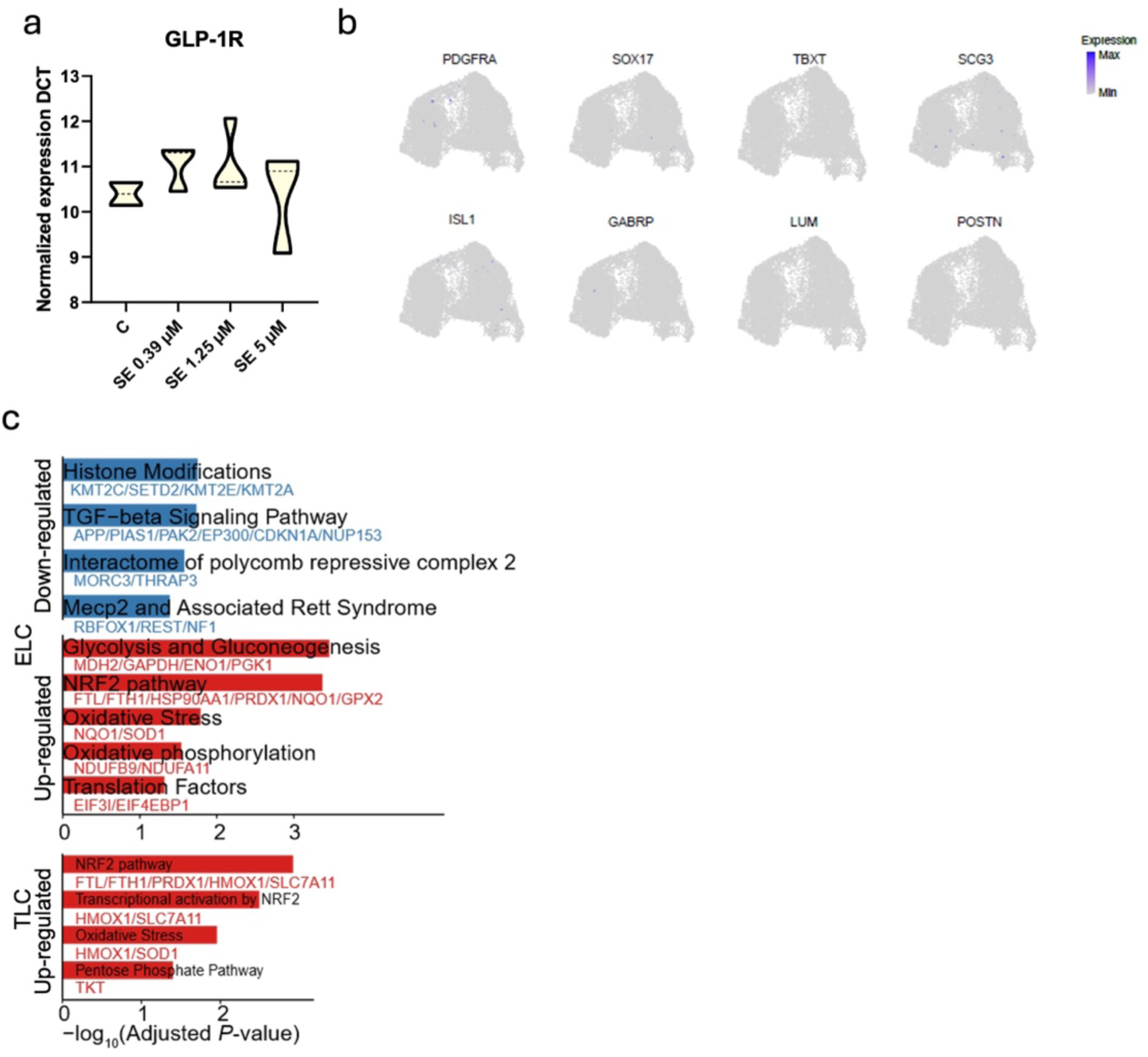
Transcriptomic analysis of blastoids treated with semaglutide (SE). **a**, *GLP-1R* mRNA expression measured by qPCR in blastoids untreated (control, C) or treated with SE (0.039 μM, 1.25 μM, or 5 μM). The experiment was conducted with four technical replicates. **b**, UMAP projection of single-cell RNA-seq data showing expression of hypoblast lineage markers (*PDGFRA, SOX17, TBXT, SCG3, ISL1, GABRP, LUM,* and *POSTN*) in blastoids. **c**, Bar plot showing up- and downregulated pathways and their consistently differentially expressed genes in SE-treated ELC and TLC 0.039 μM (L) and 5 μM (H) versus SE-untreated controls.

**Extended Data Table 1.** Differentially expressed receptivity-associated genes in endometrial epithelial organoids (EEOs).

**Extended Data Table 2.** Differentially expressed genes with and without semaglutide treatment in hormonally stimulated endometrial epithelial organoids (EEOs).

**Extended Data Table 3.** Differentially expressed gene in endometrial stromal cells after semaglutide treatment (1.25 μM and 5 μM).

**Extended Data Table 4.** Lineage-specific differentially altered metabolic activity predicted from scRNA-seq of blastoids treated with 0.039 μM (L) and 5 μM (H) semaglutide compared to control (C).

**Extended Data Table 5.** Clinical and demographic characteristics of healthy fertile women.

## Data availability

The bulk-RNA seq and single-cell RNA-seq raw data is available under accession number PRJNA1449370.

## Acknowledgements

We would like to thank all the volunteers for donating the study samples. We are also grateful to the Center for Molecular Medicine at Karolinska Institutet, the Seahorse Core Facility team, and especially Dr. Matthew Hunt for their valuable support and Annikki Löfhjelm for the excellent technical assistance in performing immunoassays. We thank Dr. Shivam Chandel and Eda Erbil for providing pancreatic β-islet organoids. This study was partially performed at the Live Cell Imaging Core facility/Nikon Centre of Excellence, at Karolinska Institutet, Sweden, supported by the KI infrastructure council.

## Fundings

The present study was funded by: Horizon Europe grant (NESTOR, grant no. 101120075) of the European Commission; Novo Nordisk Foundation (grant no. NNF24OC0092384); Estonian Research Council (grants nos. PSG1082, PRG1076); Swedish Research Council (grant no. 2024-02530); Estonian Ministry of Education and Research Centres of Excellence grant TK214 name of CoE and the Sigrid Jusélius Foundation.

## Author’s contributions

A.A., A.D.S.P., and A.S.L. initiated and established the cellular models for the endometrium, performed and participated in all experiments, analyzed the data, and wrote the manuscript. D.L. performed cAMP assays, resazurin assays, and prolactin measurements, drafted the manuscript, and contributed to the discussion. C.Z. analyzed the scRNA-seq data and contributed to the interpretation and discussion. K.K. performed endometrial immunostainings and quantifications. L.B.R. participated in the cAMP sensor experiments. S.R.D. contributed to the clinical aspects of the study. S.R. generated blastoids for scRNA-seq and performed immunostaining and edited the manuscript. I.R. and P.D. contributed to mblastoid generation for the implantation assay, edited and critically revised the manuscript. M.S. and M.P. were responsible for tissue processing and collection of endometrial samples. H.K. performed FRT analysis for glycodelin and hCG detection in the implantation model. P.D., G.A. and F.L. provided infrastructure and ensured ethical compliance for the use of blastoids and edited the manuscript. M.Z.E. participated in the data analysis and interpretation. A.S.-L. and A.S. contributed to funding acquisition, study supervision, data interpretation, and manuscript revision. A.S. provided essential infrastructure, resources and funding. All authors revised the final version of the manuscript.

## Notes

### Competing Interest Statement

The authors have declared no competing interest.

### Author Declarations

Endometrial biopsies of healthy fertile women were collected in South Estonian Hospital (Voru, Estonia) under ethical permissions No. 330/M-8 (16.10.2020) and No. 364/M-9 (16.05.2022) issued by the Research Ethics Committee of the University of Tartu. Ethical permits (454/02 and 2011/745/31-3) issued by Stockholm Region Ethical Review Authority allowed the use of embryonic stem cells to investigate human reproduction and blastoid generation.

### Summary of Updates

The version of the manuscript has been revised and few additional analysis have been added.

